# Mechanistic Insights Into Postoperative Delirium Using Untargeted High-Throughput Proteomics in Elderly Patients - A Case-Control Study

**DOI:** 10.1101/2025.05.05.25326969

**Authors:** Mario Lamping, Maria Heinrich, Vadim Farztdinov, Clarissa von Haefen, Jayanth Sreekanth, Michael Mülleder, Markus Ralser, Georg Winterer, Claudia D. Spies

**Affiliations:** Department of Anaesthesiology and Intensive Care Medicine (CCM, CVK), Charité - Universitätsmedizin Berlin, Augustenburger Platz 1, 13353 Berlin, corporate member of Freie Universität Berlin, Humboldt-Universität zu Berlin, and Berlin Institute of Health, Germany; Berlin Institute of Health (BIH), Anna-Louisa-Karsch-Straße 2, 10178 Berlin, Germany; High-Throughput Mass Spectrometry Core Facility, Charité - Universitätsmedizin Berlin, Charitéplatz 1, 10117 Berlin, corporate member of Freie Universität Berlin, Humboldt-Universität zu Berlin, and Berlin Institute of Health, Germany; Department of Biochemistry, Charité - Universitätsmedizin Berlin, Charitéplatz 1, 10117 Berlin, corporate member of Freie Universität Berlin, Humboldt-Universität zu Berlin, and Berlin Institute of Health, Germany; PI Health Solutions GmbH, Berlin, Germany; Charité - Universitätsmedizin Berlin, Charitéplatz 1, 10117 Berlin, corporate member of Freie Universität Berlin, Humboldt-Universität zu Berlin, and Berlin Institute of Health, Germany

## Abstract

**Background:** Postoperative delirium (POD) in elderly patients is a common and serious complication after surgery with an unclear pathogenesis at the molecular level. Perioperative untargeted high-throughput proteomic profiling may provide insights into underlying mechanistic molecular patterns and help identify patients at high risk, guiding preventative and therapeutic measures.

**Methods:** This study is a monocentric substudy of the European BioCog project, a prospective multicentre observational study involving elderly patients aged ≥65 undergoing elective surgery. Patients with preexisting cognitive impairment (Mini-Mental State Examination score ≤23) were excluded. POD was assessed twice daily for up to seven days using the Nursing Delirium Screening Scale (Nu-DESC) and the Confusion Assessment Method for the Intensive Care Unit (CAM-ICU), along with patient chart reviews. Proteomic profiling was conducted using high-throughput liquid chromatography mass spectrometry on sequential pre- and postoperative plasma samples. Data were analysed using a matched case-control design, employing both cross-sectional and longitudinal approaches, along with pathway enrichment analysis as a functional approach.

**Results:** A total of 226 highly abundant proteins were investigated in 168 patients (50% POD incidence). Multiple pathways, particularly those involved in the activation of the innate immune response and the complement system, were associated with POD in both cross-sectional and longitudinal analyses. Butyrylcholinesterase showed the most robust regulation, with preoperative downregulation and postoperative upregulation in patients with POD, while further downregulated in those without POD. Enzyme activity showed significant decrease in both groups. Additionally, a set of eight preoperative proteins distinguished between patients with and without POD with 86% sensitivity and 79% specificity.

**Conclusion:** Untargeted high-throughput proteomics is a feasible approach to characterize pathways involved in POD pathogenesis. This case-control study identified a protein signature associated with POD, emphasizing the need for larger cohorts to confirm these observations and improve the mechanistic understanding of POD.

**Highlights:**

- The complex pathophysiology of postoperative delirium is poorly understood
- High-throughput proteomics uncovered distinct regulated features in delirium patients
- Enrichment analysis showed differential regulation of pathways in multiple domains
- Logistic regression separates delirium patients using 8 proteins with 86% sensitivity

## Introduction

Postoperative delirium (POD) is a serious complication after surgery and anaesthesia^1^. Incidence rates vary considerably across studies, ranging from 5.1% up to 53.3% in a meta-analysis of 26 studies^1,2^, with highest incidences among elderly patients^3^. POD is related to increased morbidity and mortality, loss of autonomy, health care dependency, reduced quality of life, long-term neurocognitive disorders (NCD), depression and posttraumatic stress disorder^1^. Additionally, immense healthcare expenses result in significant socioeconomic effects^4^. Predisposing factors are numerous and include age, frailty, cognitive impairment and organ dysfunction, leaving the patient vulnerable to precipitating factors^5^. Among others, these include anaesthesia related factors (e.g. EEG burst suppression^6,7^, drugs with anticholinergic side effects), surgical trauma, duration of anaesthesia and preoperative fasting^1,8^. Suspected pathomechanisms are multifactorial and remain poorly understood. Molecular mechanisms are hypothesised to involve neuroinflammation, neurotransmitter and neuroendocrine dysregulation, network disconnectivity, mitochondrial and metabolic dysfunction, oxidative stress, autophagy, cellular aging and circadian rhythm disruption^9,10^. We postulate that POD arises from multiple etiologies linked to acute encephalopathies triggered in predisposed patients, resulting in an “acute on chronic” disease model^5,8^. We propose these to be categorised in 4 domains:

i. **toxicity axis**: anaesthesia-related toxicity, anticholinergic perioperative medication
ii. **inflammatory axis**: immune and complement activation, blood-brain barrier (BBB) disruption and neuroinflammation, endothelial dysfunction
iii. **metabolic dysregulation axis**: energy metabolism, mitochondrial dysfunction, oxidative stress, autophagy imbalance
iv. **hypoxia axis**: anaemia, hypoperfusion/hypotension, haemostatic dysregulation and coagulopathy/thrombosis

Proteomics offers direct insights into (patho-)physiology underlying complex diseases such as POD, facilitating identification of therapeutic targets and associated markers and signatures^11,12^. Multi-protein signatures have been shown to increase sensitivity, specificity and the strength of prognostic and molecular stratification^11,13^. Recent advances in the field of high-throughput liquid chromatography and mass spectrometry (HT-LCMS) allow for a cost-effective analysis of the proteomic profile, especially for a large number of samples^13^. Several candidate proteins (e.g C-reactive protein (CRP), tumour necrosis factor-alpha (TNF-ɑ), cholinesterase (ChE) activity) have been investigated in the context of POD but showed limited specificity and have not demonstrated clinical applicability to date^8,14,15^. MS has uncovered several potential molecular alterations associated with key pathological processes in POD, including neuroinflammation, oxidative stress, and synaptic dysfunction^8,9^.

Among these, proteins such as cytokines (e.g., IL-6, IL-8, TNF-ɑ), neurotrophic factors (e.g., IGF-1), and acute-phase proteins (e.g., CRP, S100B) have been identified as promising candidates for diagnostic and prognostic applications^12,15^. However, studies are limited by unstandardised designs and methods, targeted analyses and small sample sizes^12,15,16^. This study aims to i) compare perioperative plasma proteomes of patients with and without POD, ii) explore key pathophysiological pathways, and iii) characterize preoperative proteomic profiles to enhance mechanistic understanding of POD.

## Methods

### Study Design and Population

This investigation is a secondary analysis of the BioCog project (www.biocog.eu)^17^, using an innovative HT technology to assess proteomics^13^. The study included patients aged ≥65 years undergoing elective surgery of expected duration of ≥60 minutes with a Mini-Mental State Examination (MMSE) score of ≥24 points. Screening included preoperative NCD and frailty^18^.

This study was approved by the local Ethics Committees (ref.: EA2/092/14 and 14-469) and conducted in accordance with the Declaration of Helsinki (ClinicalTrials.gov: NCT02265263). Written informed consent was obtained from each patient.

### Postoperative Delirium

POD was defined according to the 5th edition of Diagnostic and Statistical Manual of Mental Disorders (DSM-5) criteria^19^. Patients were considered delirious in case of

- ≥2 cumulative points on the Nursing Delirium Screening Scale (Nu-DESC)^20^ and/or a positive Confusion Assessment Method (CAM)^5^ score and/or
- a positive CAM for the Intensive Care Unit (CAM-ICU)^21^ score and/or
- patient chart review that showed descriptions of delirium (e.g. confused, agitated, drowsy, disorientated, delirious, antipsychotic therapy).

Delirium screening started in the recovery room and was conducted twice daily (08:00 and 19:00 ±1 hour) for up to 7 days postoperatively by a research team trained and supervised by psychiatrists and delirium experts, independent of routine hospital procedures.

### Trial Design

At the time of HT proteomic analyses, the BioCog study database was completed and blood samples were stored in the lab facilities of the Charité. We conducted proteomic analyses in a matched case-control design. Given the strong influence of duration of anaesthesia (DoA) on POD^8^, we aimed to match POD cases with a control within a ±30-minute DoA range or, if impossible, patients were matched iteratively by pairing each case with a control having the closest DoA. The removal of 45 unpaired samples (i.e. pre- and postoperative sample of the *same* patient) disrupted initial case-control matching. As age and sex can influence proteomic signatures^22^, we rematched samples over sex, age and nutritional status (MNA)^23^, resulting in further removal of 34 samples (17 patients). The final balanced set contained 336 samples (see Fig. 1 and Tabs. S1-S6).

**Fig. 1.**
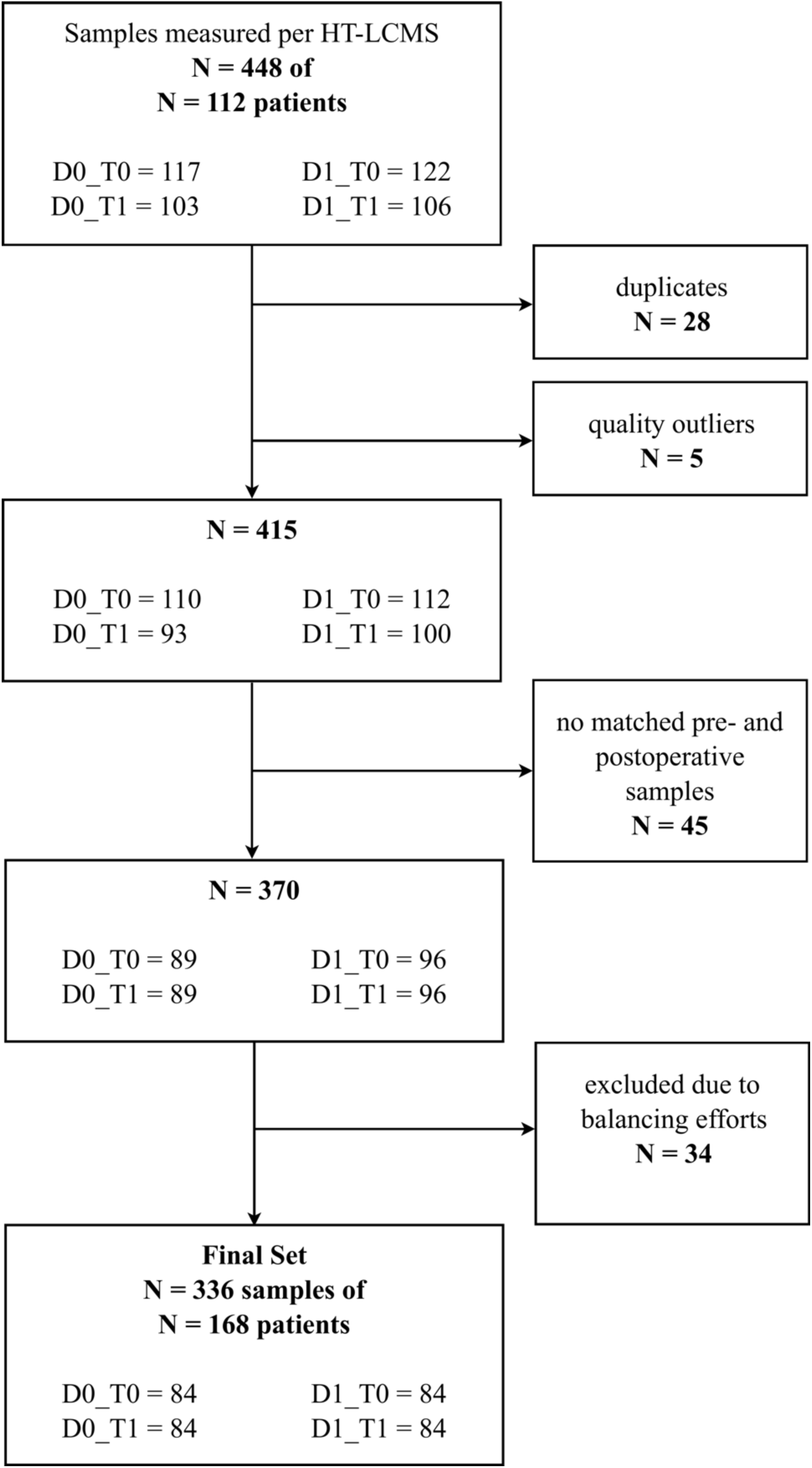
Sample Flow Chart. D0 = non-POD patients, D1 = POD patients, HT-LCMS = high throughput liquid chromatography mass spectrometry, T0 = preoperative, T1 = postoperative.

### Patient Characteristics

The patient cohort was described using physical status according to the American Society of Anaesthesiologists (ASA PS)^24^, Charlson Comorbidity Index (CCI)^25^, grade of NCD^18^ and frailty status^18^, impaired activities of daily living according to Barthel (ADL)^26^ and site of surgery.

### Blood Sampling

Blood samples were collected preoperatively on the morning of the surgery (T0) and on postoperative day 1 (T1) via venipuncture or arterial cannula. Blood samples were immediately centrifuged, plasma was extracted and frozen in 50µl aliquots at -80°C. The samples were thawed for the first time for proteomic analyses, transferred to an Eppendorf twin.tec®-PCR plate for proteomic analyses and refrozen at -80°C.

### Enzyme Activity

Free plasma butyrylcholinesterase (BCHE) activity was analysed with the point-of-care photometry “ChE Check Mobile®” (Securetec AG, Neubiberg, Germany)^27^ within one hour after sampling.

### Proteomic Analysis

To avoid batch effects and confounding of experimental and technical variables, samples were harmonised across plates according to sampling time points. Matched pairs remained together on one plate. Samples were distributed starting with the postoperative samples and then using as many time series as possible. Samples were randomised within plates. Sixteen commercial control samples (12 plasma, 4 serum) were included on each plate to detect sample preparation effects. Semi-automated sample preparation was performed in 96-well format as previously described in Messner et al.^28^, using in pre-prepared stock solution plates stored at -80°C. Briefly, 5μl of thawed plasma samples were transferred to the pre-made denaturation/reduction stock solution plates (50μl 8M Urea, 100mM ammonium bicarbonate (ABC), 5µl 5mM dithiothreitol) resuspended and incubated at 30°C for 60 minutes. 5μl were then transferred from the iodoacetamide stock solution plate (100mM) to the sample plate and incubated in the dark at room temperature for 30 minutes before dilution with 100mM ABC buffer (340μl). 220μl of this solution was transferred to the pre-made trypsin stock solution plate (12.5μl, 0.1μg/μl) and incubated at 37°C for 17 hours (Benchmark Scientific Incu-Mixer MP4). The digestion was quenched by addition of formic acid (10% v/v, 25μl) mixture was cleaned using solid phase extraction in C18 96-well plates (BioPureSPE Macro 96-Well, 100mg PROTO C18, The Nest Group). The eluted samples were vacuum dried and reconstituted in 60μl 0.1% formic acid with shaking. Insoluble particles were removed by centrifugation and the samples transferred to a new plate. A study pool was generated from all samples. Peptides were resolved on an Agilent 1290 Infinity II (Crick laboratory) in reversed phase mode using a C18 ZORBAX Rapid Resolution High Definition (RRHD) column 2.1mm x 50mm, 1.8μm particles at a column temperature of 30°C. The eluent was directed to a TripleTOF 6600 mass spectrometer (SCIEX) equipped with IonDrive Turbo V Source (SCIEX) operating in scanning SWATH mode^29^. A linear gradient was applied which ramps from 3% B to 36% B in 5 minutes (Buffer A: 0.1% FA; Buffer B: ACN/0.1% FA) with a flow rate of 800μl/minute. For washing the column, the organic solvent was increased to 80% B in 0.5 minutes and was kept for 0.2 minutes at this composition before going back to 1% B in 0.3 minutes. 5μg of peptide was injected. The DIA/SWATH method consisted of an MS1 scan from m/z 100 to m/z 1500 (20ms accumulation time) and 25 MS2 scans (25ms accumulation time) with variable precursor isolation width, covering the mass range from m/z 450 to m/z 850. Ion source gas 1 (nebulizer gas), ion source gas 2 (heater gas) and curtain gas were set to 50, 40 and 25 respectively. The source temperature was set to 450°C and the ion spray voltage to 5500V. The study pool was repeatedly injected to monitor LC-MS performance.

### Data Preprocessing

The raw proteomics data were processed using DIA-NN 1.8^30^, using standard settings except for MS1 and MS2 resolution which were adjusted to 20 and 12ppm, respectively, and scan window radius to 6. Peptides were identified with a public plasma library^31^ for which retention times and fragmentation spectra were replaced with in silico generated values by DIA-NN. DIA-NN output data matrix of normalised precursor intensities was integrated with metadata. Five samples identified by quality control as outliers were removed, defined as samples having numbers of precursors significantly less than [median - 4.3 x MAD (median absolute deviation)]. Peptides with excessive number of missing values (>40%) were excluded from our analysis. The missing values of remaining peptides were imputed group-wise (DEk_Tn) using the Principal Component Analysis (PCA) method^32^. After imputation, normalisation of the total dataset was performed using the LIMMA^33^ implementation of cyclic loess method^34^ with option *fast*^35^. It was followed by batch correction using LIMMA^33^. To obtain a quantitative protein data matrix, the log2-intensities of peptides were filtered, only peptides belonging to one protein group were kept, and then summarised into protein log intensity using the “linear models for panel data” method (PLM)^36^ implemented in the preprocessCore R package^37^.

### Statistical Analyses

We took a longitudinal and a cross-sectional approach, both using LIMMA^33^. POD patients are noted as “DE1”, non-POD patients as “DE0”. T0 refers to preoperative, T1 to postoperative day 1. The longitudinal approach used log2 ratios of protein levels at T1 relative to T0 within each group, while the cross-sectional approach analyzed log2 protein expression at individual time points, comparing groups at T0 and T1.

### Longitudinal approach

Applied model log2[*p*(T1)/*p*(T0)] ∼ 0+Class

- Contrast 1 – operation effect on non-POD patients – Class = (DE0|T1 – T0)
- Contrast 2 – operation effect on POD patients – Class = (DE1|T1 – T0)
- Contrast 3 – interaction between operation and POD (“pure” POD effect) - (DE1 – DE0|T1 – T0)

In the longitudinal approach we consider log2 ratios to baseline (T0). Therefore, all factors that do not change over time (such as genetic predispositions or consistent baseline protein expression) are canceled out. This provides higher accuracy as compared to the cross-sectional approach.

### Cross-sectional approach

Applied model log2[*p*] ∼ 0+SubClass

- Contrast 4 – difference between POD and non-POD patients before operation – (DE1 – DE0|T0)
- Contrast 5 – difference between POD and non-POD patients the day after operation – (DE1 – DE0|T1)
- Contrast 6 – interaction between operation and POD, (“pure” POD effect) – (DE1 – DE0|T1 – T0)

Baseline characteristics are expressed as median (including lower and upper quartile) or as mean (± standard deviation, SD), except for categorical data, which are expressed as frequencies. Differences between groups were tested using Mann-Whitney U test or χ2 test. Regulated proteins were described in foldchanges (FC) and considered significant in case of a FC of 1.1 (≙log2(1.1)=0.1375) and significance level of p<0.05 (≙-log10(0.05)=1.30103). FCs are presented in log2(FC), p-values in -log10(p). Statistical analysis of BCHE enzyme activity was performed using the Wilcoxon signed-rank test due to non-normal data distribution (SPSS Statistics, version 30, ©1989, 2024 by SPSS Inc., Chicago, Illinois, USA). A p-value of <0.05 was considered significant.

### Functional Analyses

Gene set enrichment analyses (GSEA) were performed using the clusterProfiler R package^38^ with input derived from statistical contrasts (see above). Results were based on the Reactome Pathway Database (https://reactome.org), applying the Benjamini-Hochberg method for false discovery rate (FDR) correction^39^. To ensure comprehensive coverage, the Gene Ontology Biological Process (GOBP, https://geneontology.org) database was also used, with results included in the Supplementary Material. Pathway enrichment was assessed using the normalised enrichment score (NES).

### Exploratory Proteomic Classification Analysis

Logistic regression (LR) using a generalised linear model from R package caret^40^ was applied to classify patients based on preoperative proteomic profiles. Protein selection was based on LIMMA results for Contrast 4 (α=0.142 and log2(FC) ≥0.1), initially identifying 32 proteins. These candidates were further reduced using recursive feature elimination to maximise receiver operating characteristic (ROC) area under the curve (AUC). The final analysis was based on 8 proteins and employed 5 repeats of 12-fold cross-validation. ROC, AUC, sensitivity, specificity and accuracy were calculated.

### Results

In this matched case-control study we investigated the pre- and postoperative proteomic profile in 168 patients with a 50% POD incidence. Patients were balanced in terms of sex, age, MNA, preexisting NCD and site of surgery. Despite efforts to balance for DoA, there was a significant difference regarding the latter (Tabs. 1, S5 and S6). POD patients showed significantly higher ASA PS, CCI scores, impaired daily activities and frailty (Tab. 1). We identified a total of 226 proteins that were reliably measured in at least 60% of samples in each group.

**Tab. 1.**
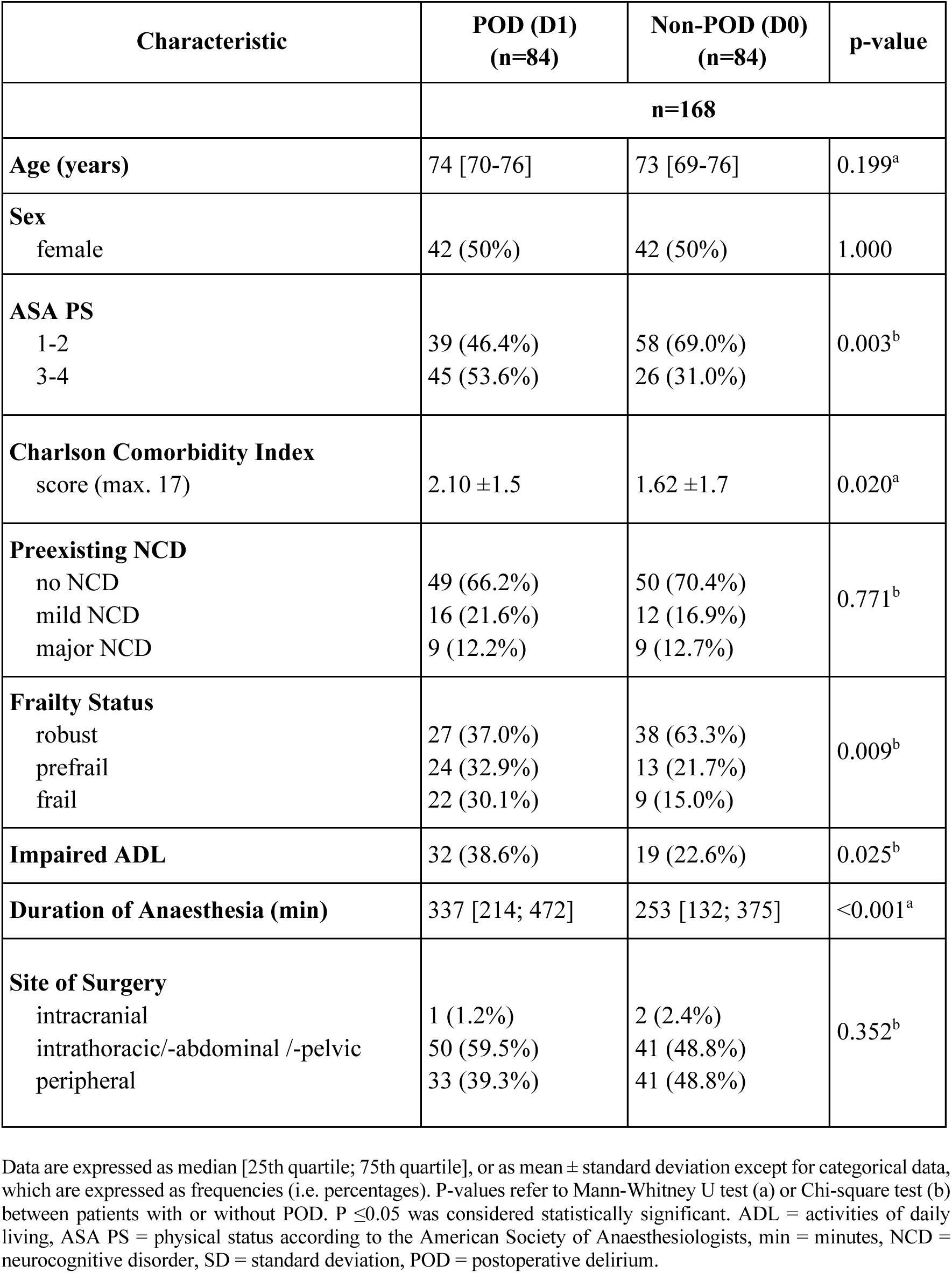
Patient Characteristics.

| Characteristic | POD (D1)<br>(n=84) | Non-POD (D0)<br>(n=84) | p-value |
| --- | --- | --- | --- |
|  | **n=168** |  |  |
| **Age (years)** | 74 [70-76] | 73 [69-76] | 0.199<sup>a</sup> |
| **Sex**<br>female | 42 (50%) | 42 (50%) | 1.000 |
| **ASA PS**<br>1-2<br>3-4 | 39 (46.4%)<br>45 (53.6%) | 58 (69.0%)<br>26 (31.0%) | 0.003<sup>b</sup> |
| **Charlson Comorbidity Index**<br>score (max. 17) | 2.10 ±1.5 | 1.62 ±1.7 | 0.020<sup>a</sup> |
| **Preexisting NCD**<br>no NCD<br>mild NCD<br>major NCD | 49 (66.2%)<br>16 (21.6%)<br>9 (12.2%) | 50 (70.4%)<br>12 (16.9%)<br>9 (12.7%) | 0.771<sup>b</sup> |
| **Frailty Status**<br>robust<br>prefrail<br>frail | 27 (37.0%)<br>24 (32.9%)<br>22 (30.1%) | 38 (63.3%)<br>13 (21.7%)<br>9 (15.0%) | 0.009<sup>b</sup> |
| **Impaired ADL** | 32 (38.6%) | 19 (22.6%) | 0.025<sup>b</sup> |
| **Duration of Anaesthesia (min)** | 337 [214; 472] | 253 [132; 375] | <0.001<sup>a</sup> |
| **Site of Surgery**<br>intracranial<br>intrathoracic/-abdominal /-pelvic<br>peripheral | 1 (1.2%)<br>50 (59.5%)<br>33 (39.3%) | 2 (2.4%)<br>41 (48.8%)<br>41 (48.8%) | 0.352<sup>b</sup> |

Data are expressed as median [25th quartile; 75th quartile], or as mean ± standard deviation except for categorical data, which are expressed as frequencies (i.e. percentages). P-values refer to Mann-Whitney U test (a) or Chi-square test (b) between patients with or without POD. $P \leq 0.05$ was considered statistically significant. ADL = activities of daily living, ASA PS = physical status according to the American Society of Anaesthesiologists, min = minutes, NCD = neurocognitive disorder, SD = standard deviation, POD = postoperative delirium.

### Statistical Analyses

#### Longitudinal Approach

In the longitudinal approach, we found 45 proteins to be regulated from pre- to postoperative in non-POD patients (Contrast 1) and 42 proteins in POD patients (Contrast 2, Figs. 2a and 2b). Fifteen proteins are regulated in non-POD patients only (Contrast 1), 8 of which are immunoglobulins. Fifteen proteins are regulated in POD patients only (Contrast 2). Among them QSOX1, GPX3, MBL2, CFHR2, C1R, TMSB4X are upregulated, and PGLYRP2, CNDP1, SERPIND1, APOA4, APCS, HBA1 and HBB downregulated.

**Fig. 2a.**
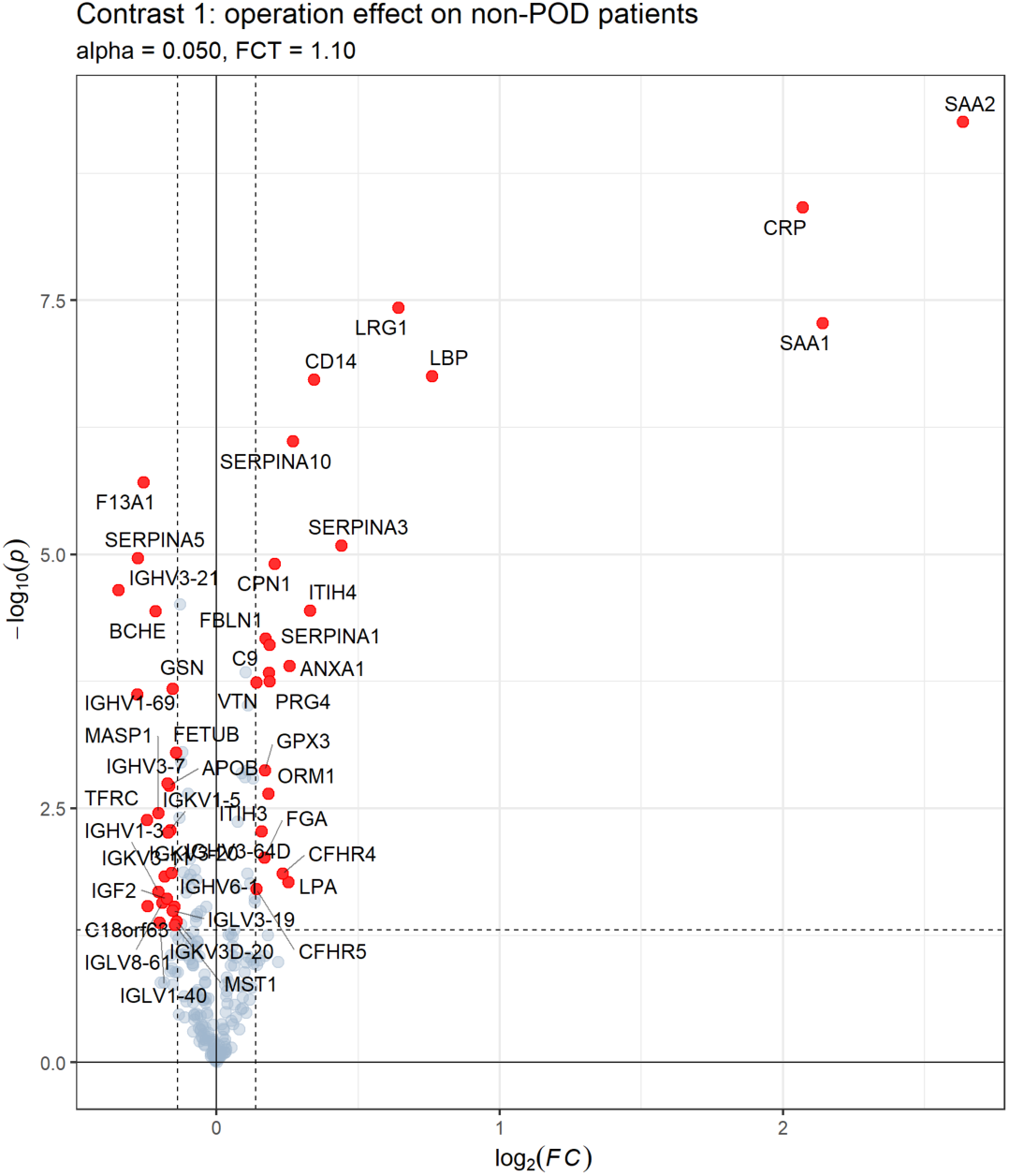
Volcano plot for contrasts of protein expression using the longitudinal approach with Contrast 1: expression profile regarding operation effect on non-POD patients. Significantly regulated proteins (p<0.05) are shown in red. Fold change (FC) threshold: 1.1, p-value is displayed in -log10(p), FC in log2(FC).

**Fig. 2b.**
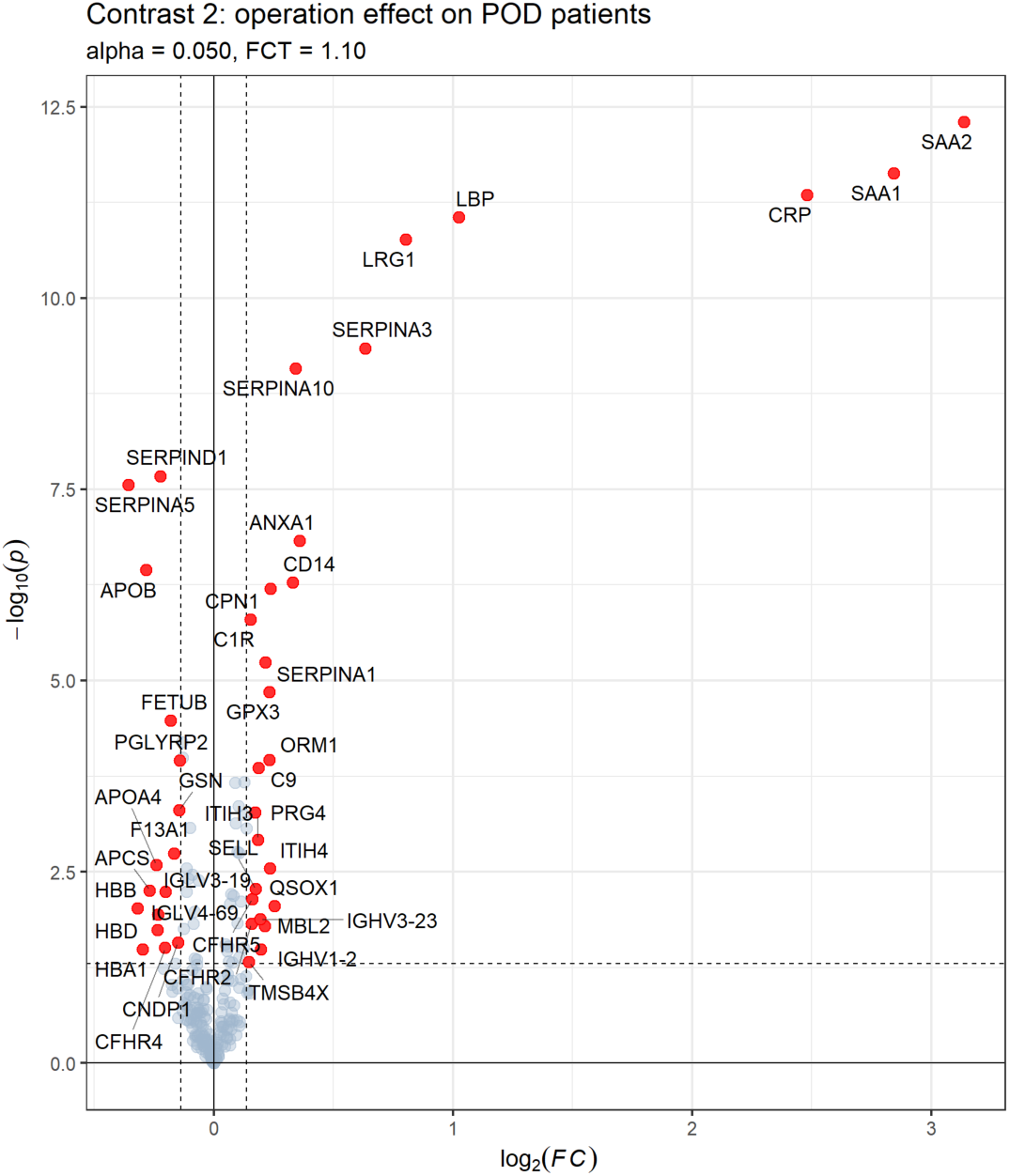
Volcano plot for contrasts of protein expression using the longitudinal approach with Contrast 2: expression profile regarding operation effect on POD patients. Significantly regulated proteins (p<0.05) are shown in red. Fold change (FC) threshold: 1.1, p-value is displayed in -log10(p), FC in log2(FC).

Of special interest are proteins regulated regarding the interaction between operation and POD (Contrast 3), also 15 in total (Fig. 2c). Most of them (except IGHV3-21 and IGHV1-69) have opposite regulation in patients with and without POD (Contrasts 1 and 2), respectively. One protein, CFHR4, is regulated in all three contrasts. Two proteins, SELL, and HBD are significantly regulated only in Contrasts 2 and 3 (SELL is up-regulated in POD patients, while HBD is down-regulated).

**Fig. 2c.**
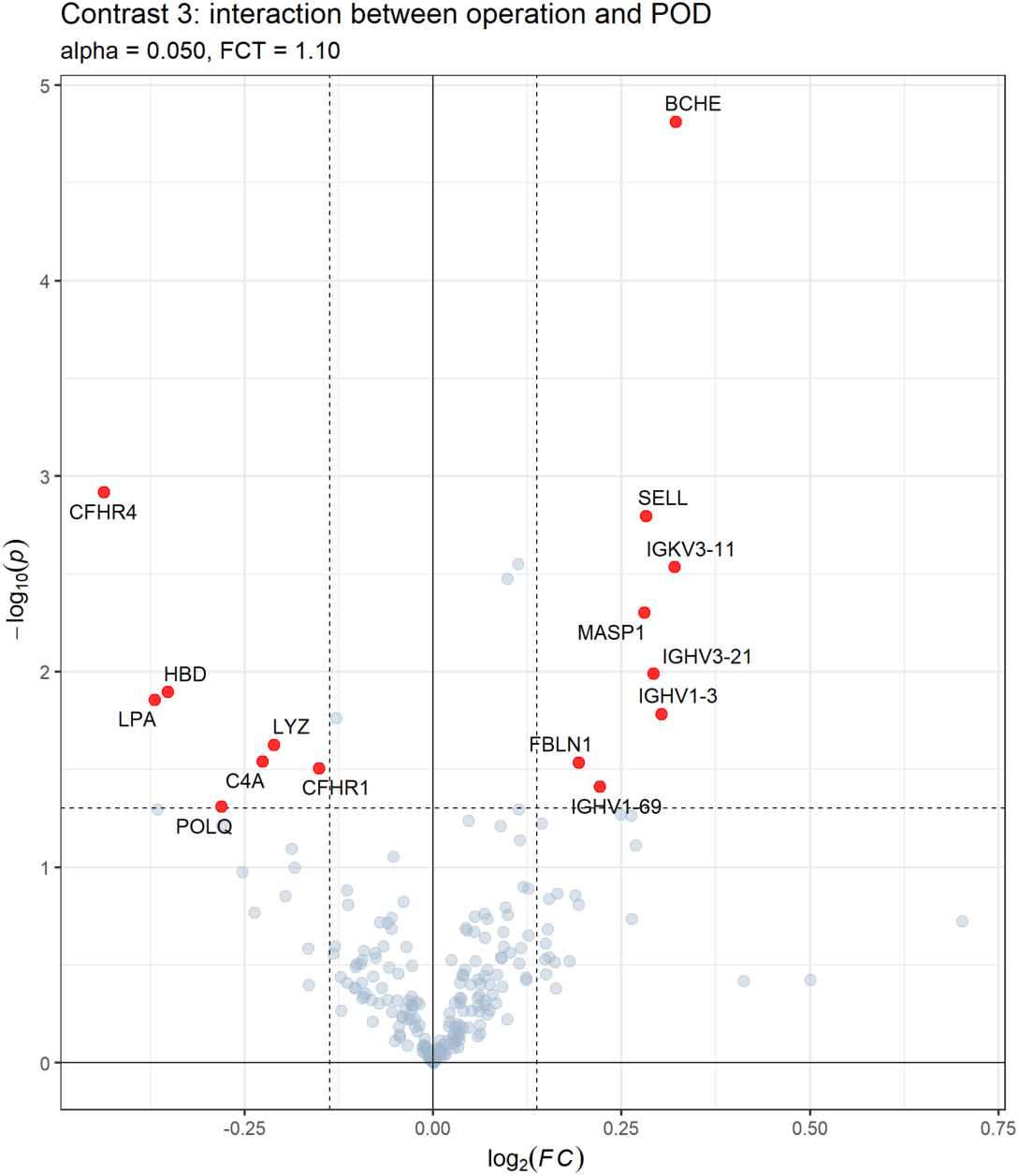
Volcano plot for contrasts of protein expression using the longitudinal approach with Contrast 3: expression profile regarding interaction between operation and POD (pure POD effect). Significantly regulated proteins (p<0.05) are shown in red. Fold change (FC) threshold: 1.1, p-value is displayed in -log10(p), FC in log2(FC).

A subgroup of interest is regulated in Contrast 1 and 3. While BCHE, MASP1, IGKV3-11, IGHV1-3, IGHV3-21 and IGHV1-69 are downregulated and LPA is upregulated in Contrast 1, they all show opposite regulations in Contrast 3, indicating significance for POD development (see Fig. S1 for scatterplot).

#### Cross-sectional Approach

The cross-sectional analysis shows a weak POD effect at T0, with Fig. 3a showing only 16 significantly regulated proteins in Contrast 4. The most significant are BCHE (log2(FC)∼0.3; FC∼1.23) and beta-2-microglobulin (B2M, log2(FC)∼0.275; FC∼1.21). The effect size at T1 (Contrast 5) is only marginally larger (Fig. 3b). Contrast 6 shows very moderate regulation (Fig. 3c). Scatterplots for Contrasts 4 and 5 are found in Fig. S2. BCHE, QSOX1, and IGKV3-11 are present in Contrasts 4 and 6, with lower preoperative expression in POD patients. Postoperatively, their expression increases, reaching levels similar to non-POD patients for QSOX1 and slightly higher for BCHE and IGKV3-11 (see Fig. 4a), explaining their absence in Contrast 5. While BCHE is upregulated in POD patients at T1, analysis of BCHE enzyme activity showed significant postoperative decrease in both groups. In the POD group, the median BCHE activity decreased from 2719.85 U/l (IQR: 2119.75– 3311.63; 95% CI: 2591.17–3044.91) preoperatively to 2410.46 U/l (IQR: 1737.43–3024.13, 95% CI: 2226.13–2701.90) (Wilcoxon signed-rank test: W=635.0; p<0.001, r=0.49). In non-POD patients, BCHE activity significantly decreased from 3052.45 U/l (IQR: 2560.50– 3530.13; 95% CI: 2888.73–3216.40) to 2634.40 U/l (IQR: 2102.85–3326.58; 95% CI: 2409.10–2922.20) (W=784.0, p<0.001, r=0.43). Their respective box plots are shown in Fig. 4b. Similarly, 9 proteins regulated only in Contrast 4 (e.g. S100A9, IGF2, APOD, LYZ, PROZ, B2M) show a comparable trend without a reversal in regulation. These 12 proteins collectively indicate an interaction with the operation. Five proteins (MASP1, IGHV3-21, SELL, HBD, CFHR4) overlap between Contrasts 5 and 6 (Fig. S2), 8 are regulated in Contrast 6.

**Fig. 3a.**
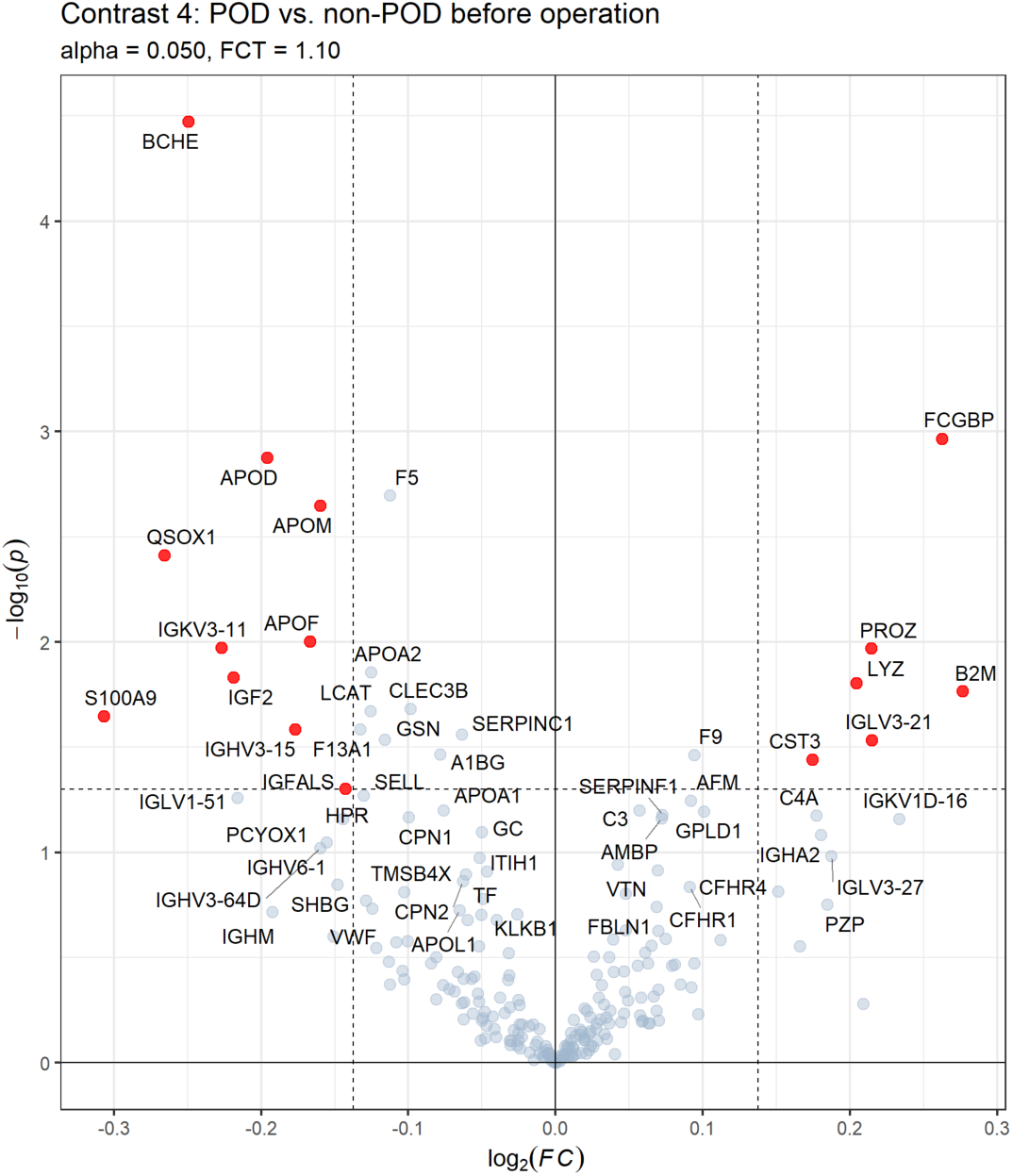
Volcano plots for contrasts of protein expression using the cross-sectional approach with Contrast 4: expression profile regarding differences between POD and non-POD patients before operation (T0). Significantly regulated proteins (p<0.05) are shown in red. Fold change (FC) threshold: 1.1, p-value is displayed in -log10(p), FC in log2(FC).

**Fig. 3b.**
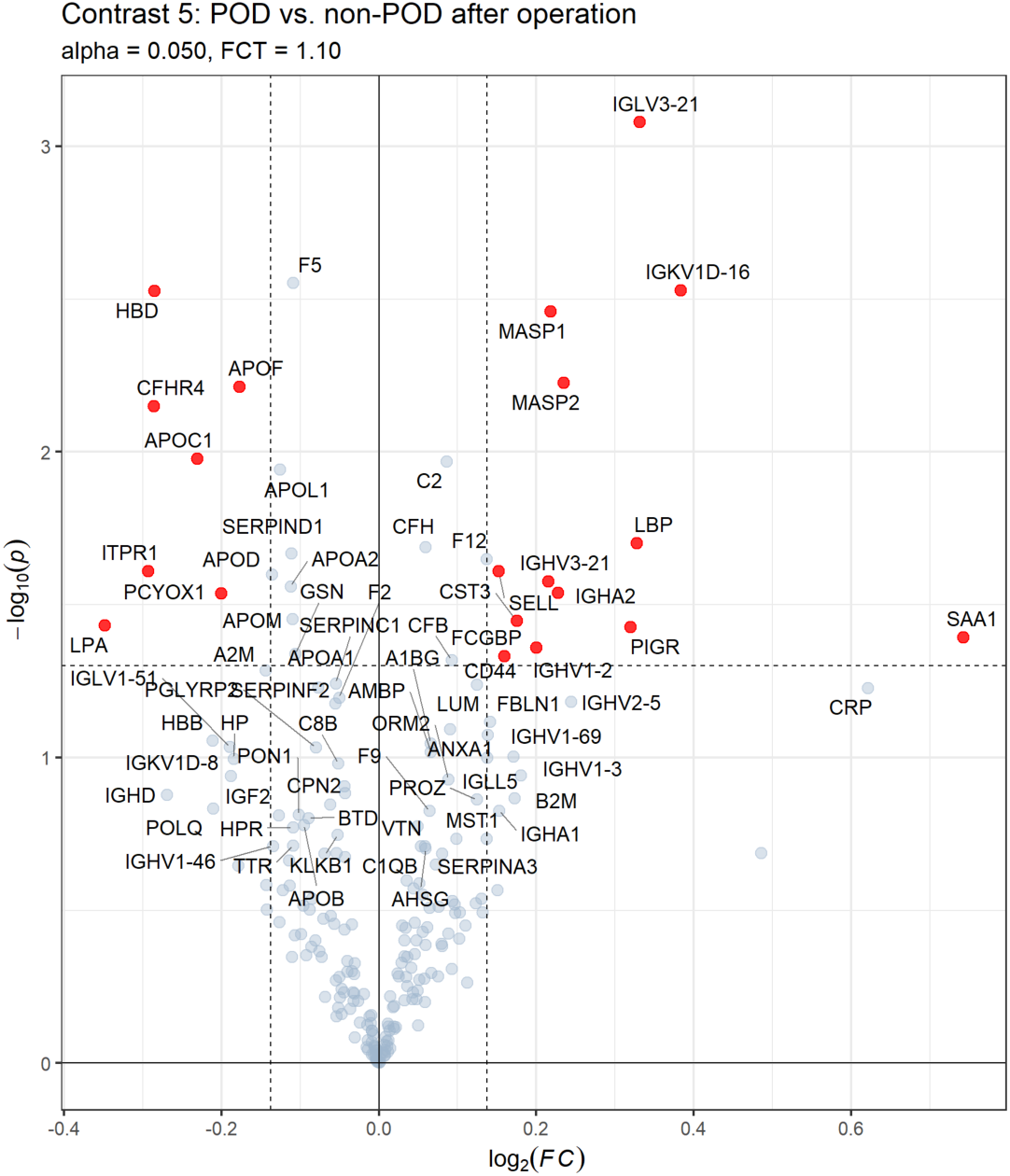
Volcano plots for contrasts of protein expression using the cross-sectional approach with Contrast 5: expression profile regarding differences between POD and non-POD patients after operation (T1). Significantly regulated proteins (p<0.05) are shown in red. Fold change (FC) threshold: 1.1, p-value is displayed in -log10(p), FC in log2(FC).

**Fig. 3c.**
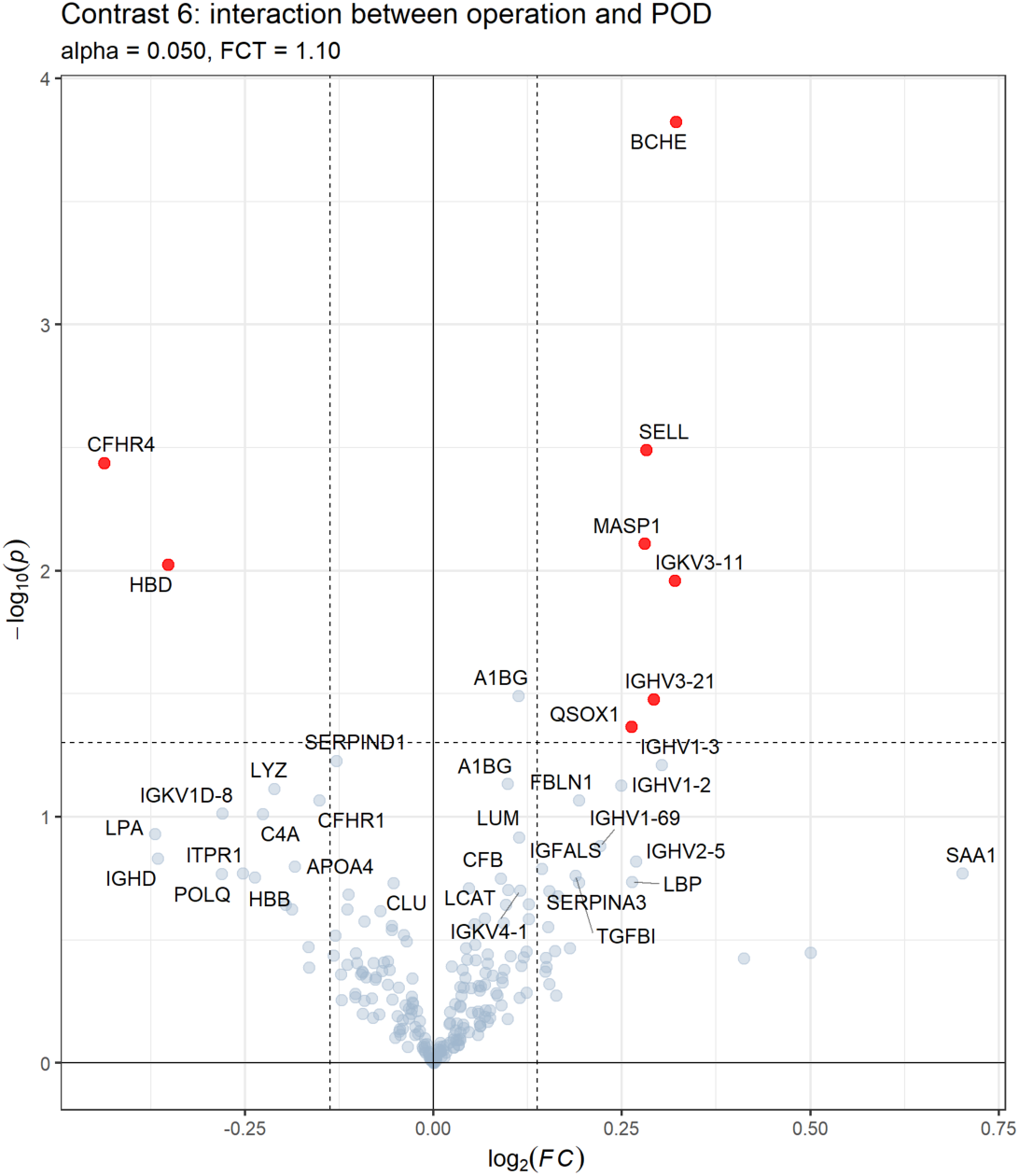
Volcano plots for contrasts of protein expression using the cross-sectional approach with Contrast 6: expression profile regarding interaction between operation and POD (pure POD effect). Significantly regulated proteins (p<0.05) are shown in red. Fold change (FC) threshold: 1.1, p-value is displayed in -log10(p), FC in log2(FC).

**Fig. 4a.**
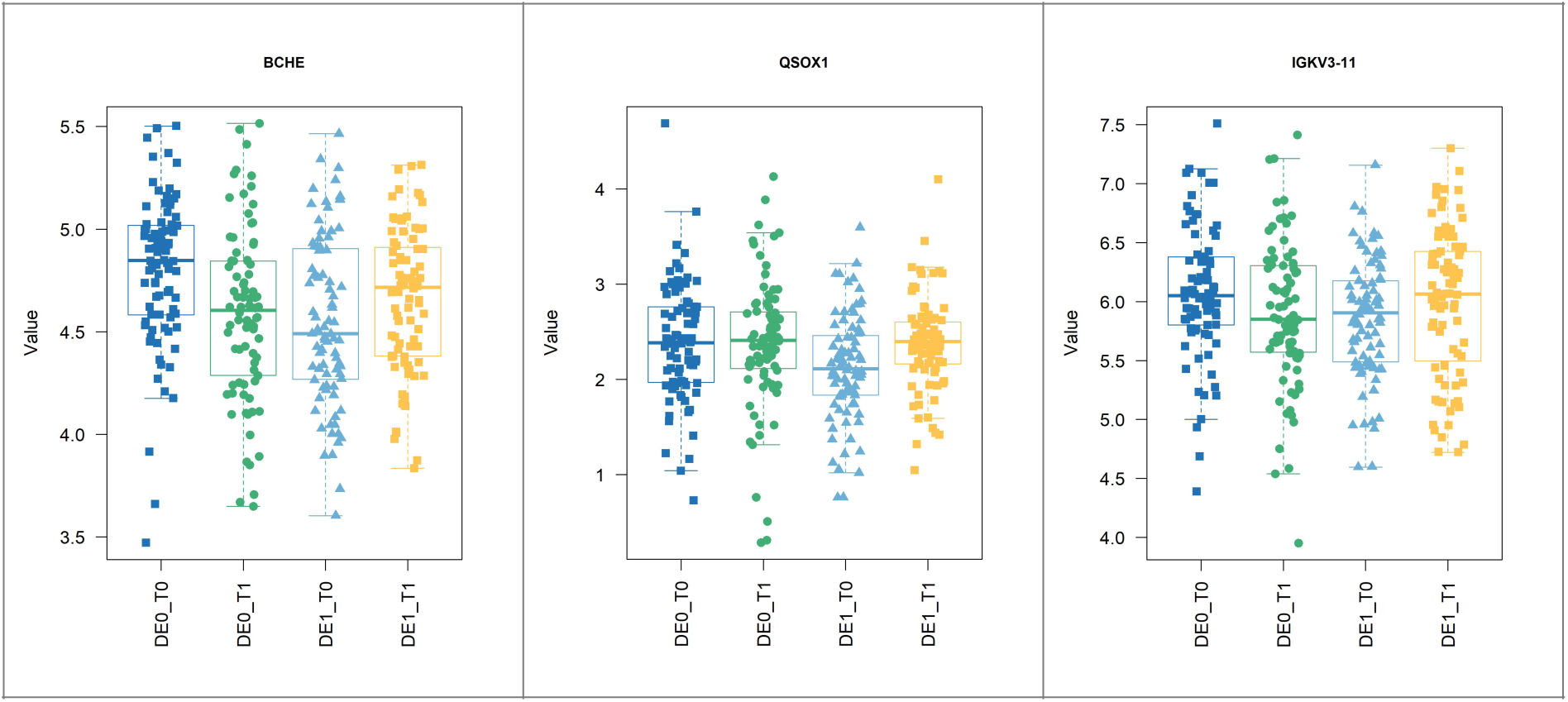
Box plots of BCHE, QSOX1 and IGKV3-11 showing preoperative (T0) and postoperative (T1) expression levels (value = log2(Int)) in non-POD (DE0) and POD (DE1) patients.

**Fig. 4b.**
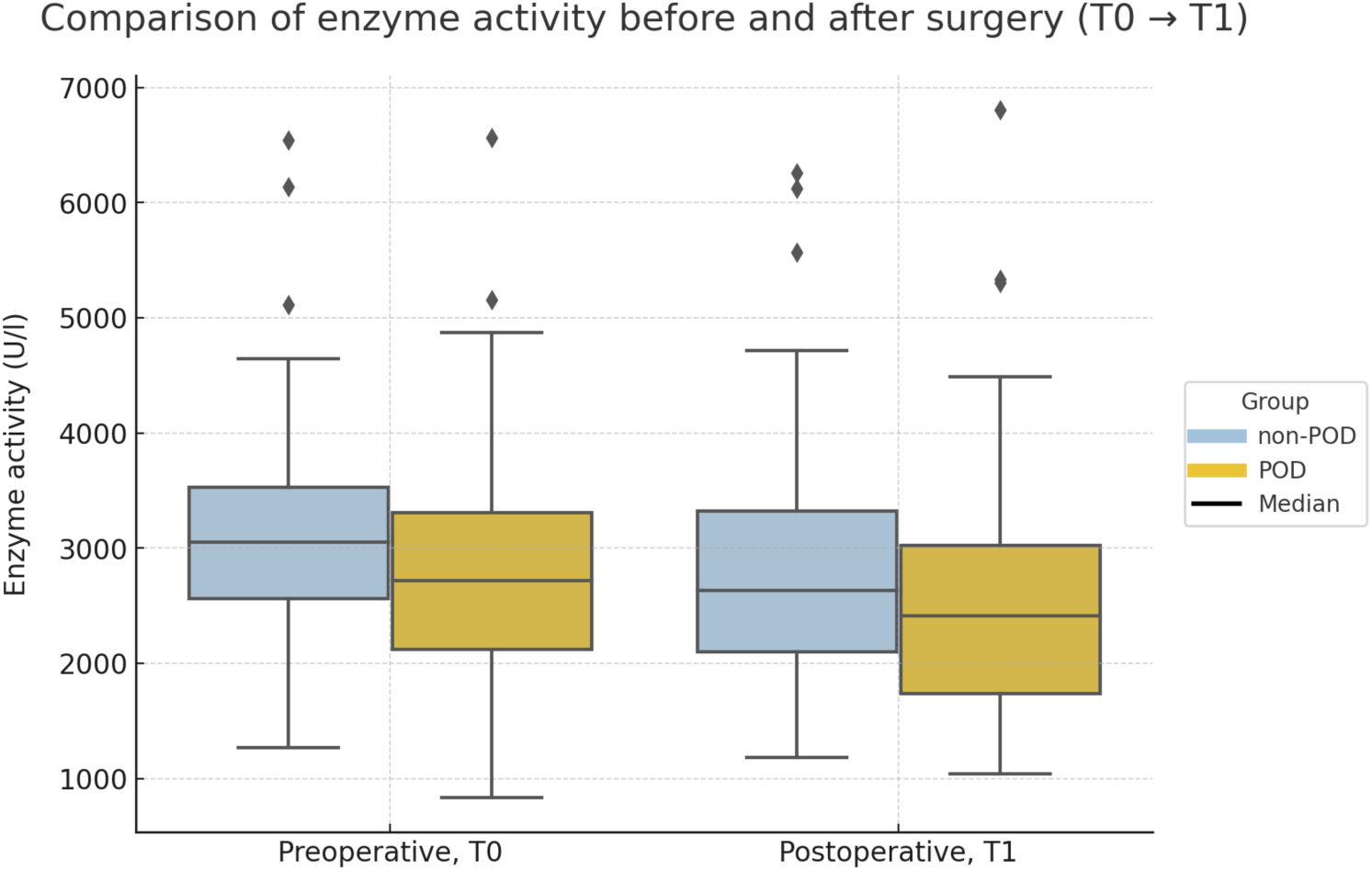
Box plots showing butyrylcholinesterase activity (U/l before (T0) and after (T1) surgery in non-POD and POD patients.

### Functional Analyses

The GSEA results for Reactome are exploratory and complex in nature, here we highlight our key findings.

#### Longitudinal Approach

Fig. 5 shows longitudinal contrasts, highlighting the operation effect. Non-POD patients generally exhibit more significant results by FDR. Among 32 inflammatory/immune-related pathways, we observe complex regulatory patterns. While many pathways, like *Toll-Like Receptor*-associated pathways, show significant upregulation in both cohorts, others are downregulated with a more pronounced effect in non-POD patients (i.e. higher activity in POD patients) with a change of direction in Contrast 3, such as *FCGR/FCERI*-associated pathways. Some display downregulation in healthy patients (C1) and upregulation in both Contrasts 2 and 3, such as *Adaptive Immune System* or 4 *Complement*-related pathways, indicating high activity.

**Fig. 5.**
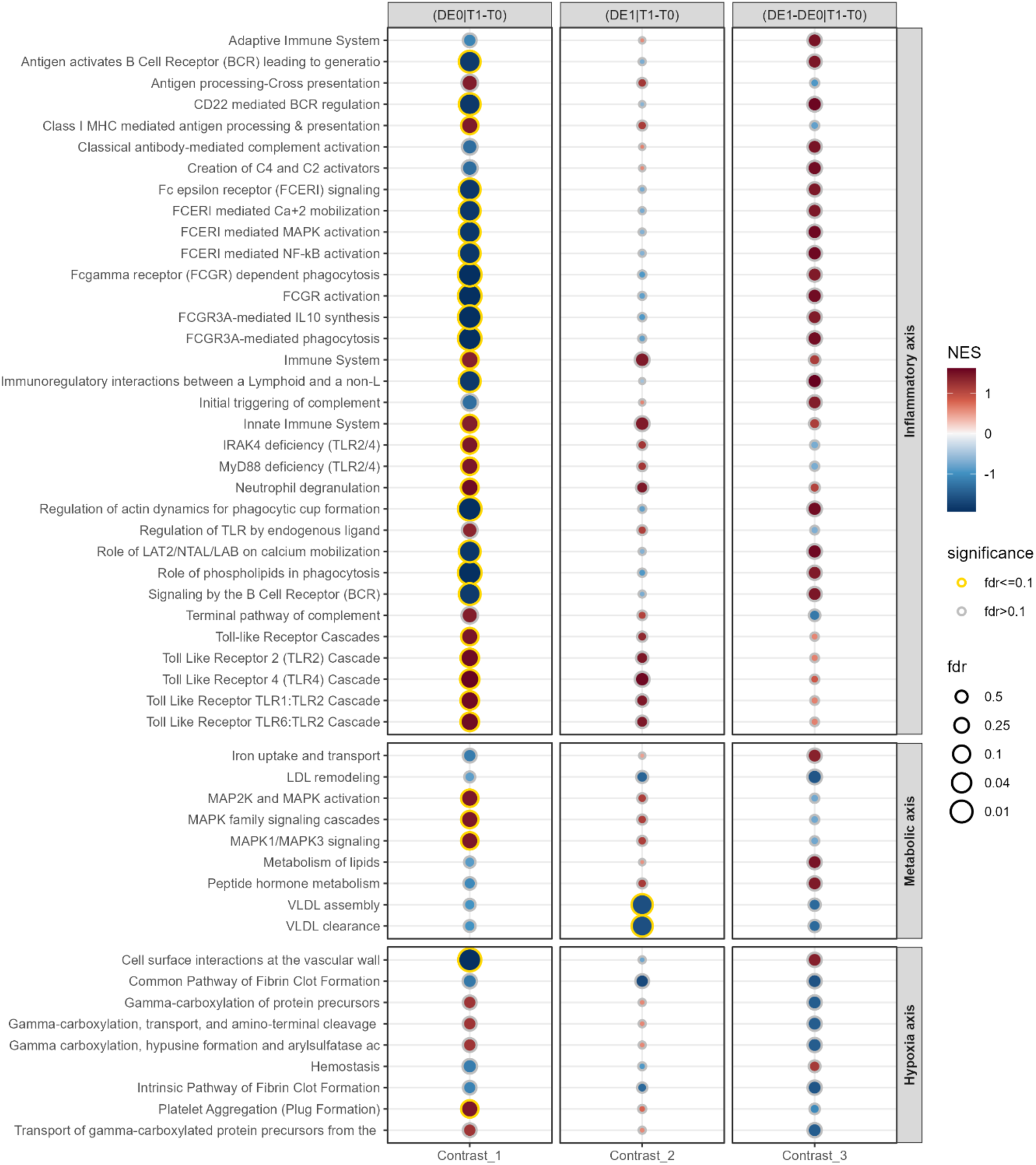
REACTOME pathway enrichment for longitudinal approach (Contrasts 1-3). Pathways with FDR ≤0.1 are marked with a golden halo. Pathways are grouped into the pathophysiology domains: Inflammatory axis, hypoxia axis and metabolic axis (see Introduction). FDR = false discovery rate, NES = normalised enrichment score. FDR is displayed in -log10(FDR).

Nine metabolic axis pathways show rather divergent behaviour. In POD patients, *Iron uptake and transport, Peptide hormone metabolism* and *Metabolism of lipids* show enrichment in Contrast 3.

Hypoxia/haemostasis axis pathways including 4 *Gamma carboxylation*-related pathways (clotting factor synthesis) and 1 (*Platelet Aggregation*) show strong or weak upregulation in Contrasts 1 and 2, respectively, and a downregulation in Contrast 3. Contrastingly, 2 *fibrin*-pathways exhibit negative enrichment across all contrasts. No toxicity axis-related pathways were observed. For further information, see Fig. S3 and Tab. S7.

### Cross-sectional Approach

Fig. 6 presents cross-sectional contrasts. Significance is generally stronger in Contrast 5. Out of 27 inflammatory axis pathways, 25 demonstrate postoperative enrichment in both groups, with the majority exhibiting greater upregulation in POD patients. Two *Interleukin*-related pathways show postoperative downregulation in healthy patients. Interestingly, 2 complement pathways show stronger enrichment in Contrast 4 (*Activation of C3 and C5, Regulation of Complement cascade*).

**Fig. 6.**
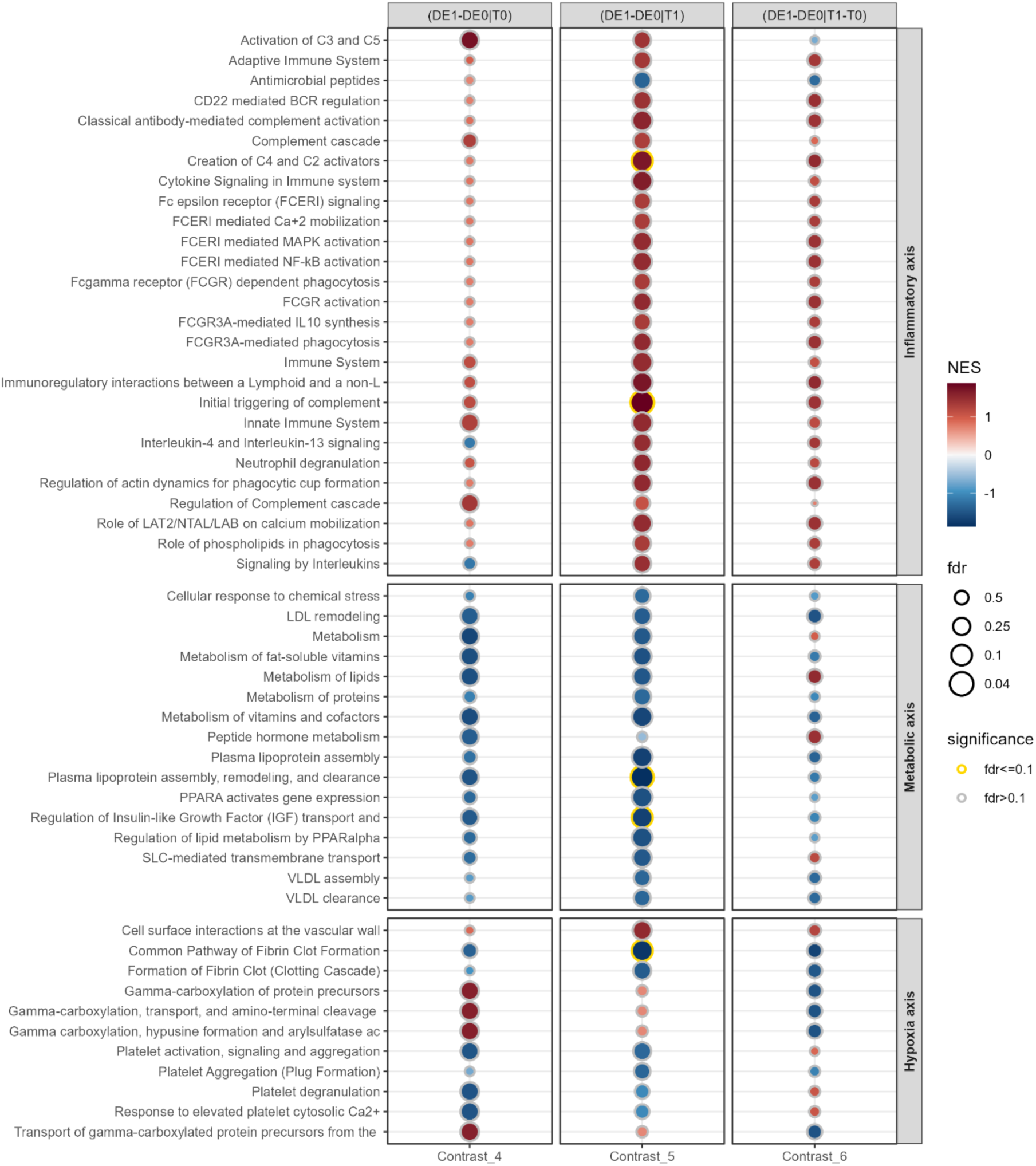
REACTOME pathway enrichment for cross-sectional approach (Contrasts 4-6). Pathways with FDR ≤0.1 are marked with a golden halo. Pathways are grouped into the pathophysiology domains: Inflammatory axis, hypoxia axis and metabolic axis (see Introduction). FDR = false discovery rate, NES = normalised enrichment score. FDR is displayed in -log10(FDR).

All 16 metabolic axis pathways show preoperative downregulation in POD patients with only some of them displaying further silencing after surgery. For *Metabolism*, *Metabolism of lipids* and *Peptide hormone metabolism*, we observe downregulation before and after surgery, but upregulation in Contrast 6.

Ten hypoxia axis related pathways show diverse enrichments: whilst *Gamma-carboxylation* pathways are highly enriched before surgery, they are silenced postoperatively. *Platelet*-related pathways show downregulation both pre- and postoperatively, but 3 show activation for pure POD effect. No toxicity axis-related pathways were observed. For further information, see Fig. S4 and Tab. S8.

### Exploratory Proteomic Classification Analysis

The LR applied to 8 protein expressions showed an AUC of 0.86 with an accuracy of 0.81 (Fig. 7a and b). Sensitivity and specificity were 0.83 and 0.79, respectively. The final list of proteins included BCHE, F5 (factor V), IGKV3-11, IGHV3-15, QSOX1, PROZ, IGLV3-27 and IGLV3-21. For a summary of their respective importance see Tab. S9.

**Fig. 7a.**
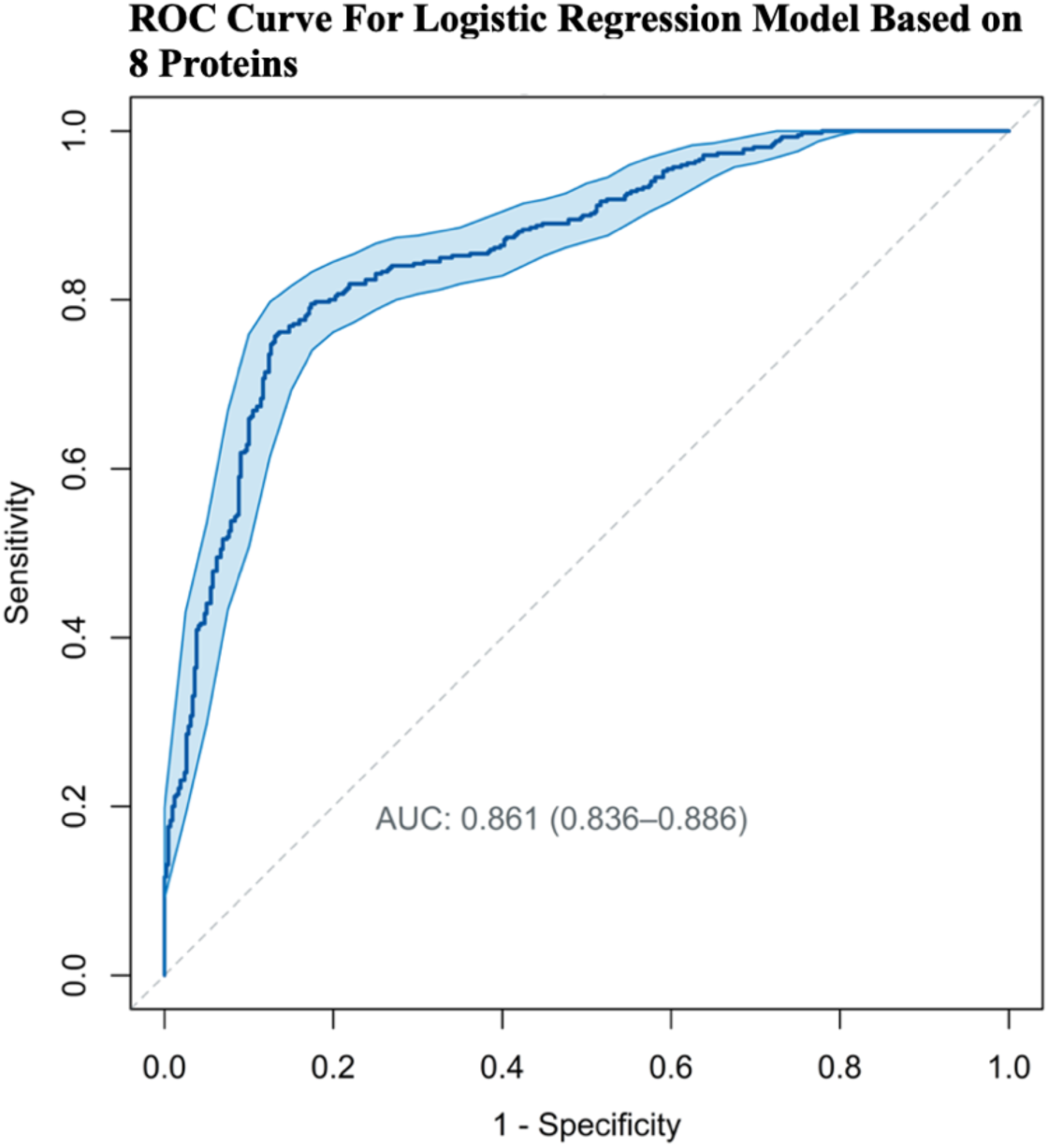
Receiver operating characteristics (ROC) of logistic regression (LR) based on preoperative expression profiles (Contrast 4, see Methods).

**Fig. 7b.**
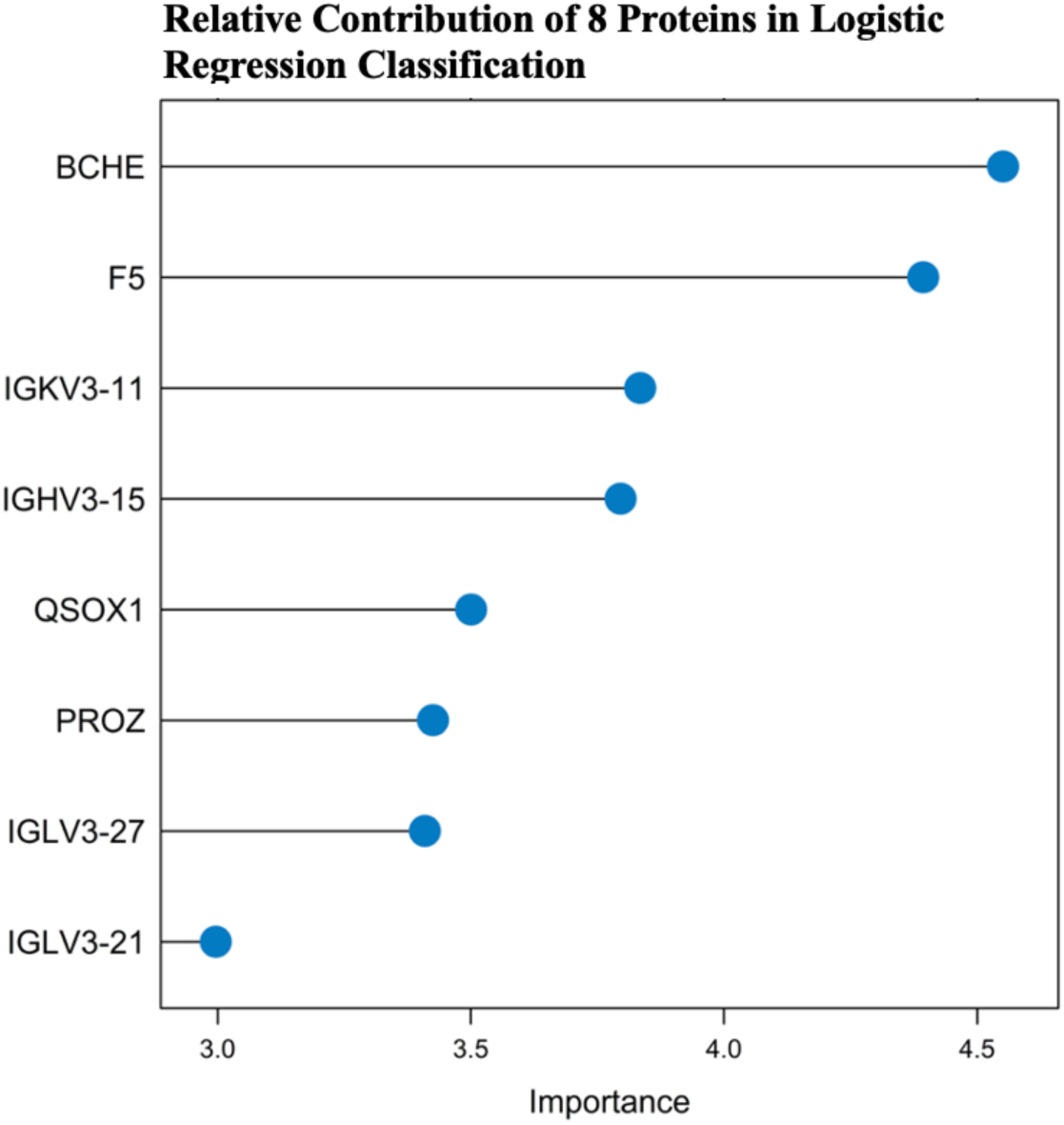
Feature contribution of the 8 most discriminative proteins identified by logistic regression (LR) based on generalised linear modelling. Importance (x-axis) is displayed based on z-values derived from LR. For further data see also Tab. S9.

## Discussion

We analysed perioperative proteomics in 168 elderly patients using HT-LCMS, identifying 226 abundant plasma proteins. POD patients had higher (pre-)frailty scores, ASA PS, and CCI classifications, underwent more intracavitary surgeries and experienced prolonged DoA. This is the first study to apply untargeted proteomics using both longitudinal and cross-sectional designs, integrated with GSEA and preoperative proteomic profiling related to POD, providing mechanistic insights into perioperative vulnerability and identifying pathophysiologic domains and perioperative alterations. This addresses key limitations of prior OMICS studies that have primarily focused on postoperative (i.e. diagnostic) features^16^. While CSF studies^41–44^ may better reflect central nervous system changes due to CSF’s proximity to the brain, plasma sampling is less invasive and may capture alterations secondary to increased BBB permeability in the elderly^16^.

### Longitudinal and Cross-sectional Approach

We observed an expected increase in acute-phase proteins, including CRP, SAA1/2, and LBP - all of which showed higher expression in POD patients. LBP has been linked to post-stroke delirium^45^, while SAA1/2 can migrate to the brain, enhance neuroinflammation and memory decline and could serve as severity markers for traumatic brain injury^46–48^. IL-6 is among the most frequently reported proteins in previous studies^16^, however, we did not detect any regulation, possibly due to the absence of pre-analytical immunodepletion. Some authors recommend this step to remove high-abundant proteins (e.g., albumin, immunoglobulins), as they can hinder the detection of low-abundant proteins in MS, which includes many POD alterations such as cytokines^16,49,50^. MASP1 (mannose-associated serine protease 1) is significantly increased in POD and downregulated in non-POD patients. It activates endothelial cells and initiates the lectin pathway of the complement system by cleaving complement C4 and C2 - a finding confirmed in the GSEA (pathway *Creation of C4 and C2 activators*, Fig. 5 and 6)^51,52^.

CFHR4 is downregulated in our POD cohort. Functional data is conflicting: mainly its inhibitory effects on the complement system were investigated, whereas new data also indicate activating functions via complex interactions^53^. Another key protein seems to be QSOX1, especially in relation to autophagy, as this process is considered in neuroinflammation^54^. It is downregulated preoperatively, showing enrichment after surgery, but only to the level of non-POD patients, who do not exhibit a significant FC (Fig 4a). QSOX1 has unclear implications. Some authors find antiapoptotic and protective effects QSOX1-expressing cells as a reaction to oxidative treatment as well as inhibition of autophagy^55,56^, others demonstrate autophagy stimulation via mitochondrial apoptosis^57^. However, upregulation of QSOX1 seems to be a reaction to oxidative stress rather than a cause^56^.

The most outstanding protein in our analysis is BCHE: it is ubiquitously found, particularly in the liver, blood, pancreas and central nervous system (glial cells)^58^. It serves as the primary acetylcholine (ACh)-hydrolyzing enzyme in human blood and contributes to inflammation and cognitive decline by inhibiting the cholinergic anti-inflammatory pathway^59^. The association of preoperative ChE activity deficits and POD has been previously reported^60–64^.

We observed a preoperative deficit and postsurgical upregulation in POD patients, while non-POD patients show a postsurgical downregulation. However, this refers to FCs (i.e. relative concentration changes). In contrast, enzymatic activity was significantly reduced in both groups post-surgery. While specific data on this phenomenon are lacking, several mechanisms could be assumed, particularly inflammation and oxidative stress^65^. Oxidation and nitrosation of cysteine residues increase disulfide bonds and formation of cystine, therefore altering protein stability and function in a powerful manner and nitrosation is caused by the inducible nitric oxide synthase (iNOS), which is induced under inflammatory conditions^66,67^. The correlation between (oxidative) stress and reduced BCHE has been shown in rats^68^, further data have shown complementary results in patients with obstructive lung disease^69^. Furthermore, isoforms of BCHE can result in different activity levels^70^. Other mechanisms of reduced activity may involve protein aggregation and inhibition with alternate substrate^71^. The POD-associated increase in protein amount may represent a compensatory mechanism aimed at mitigating preexisting ACh deficits by supplying more substrate as well as compensating for the BCHE activity decline. Of note, astrocytes upregulate BCHE expression under inflammatory conditions mediated by iron dysregulation, resulting in cognitive decline^72^.

In an analysis of a BioCog subcohort (n=127 patients with abdominal surgery, 41% POD incidence), Bosancic et al. observed similar activity (i.e., U/l) declines, although statistically not significant^64^. In the CESARO study, results were comparable to that, with a more pronounced decline in patients >70 years of age^73^. Postoperative enzyme activity declines in POD patients were also shown in a subanalysis of our DEXDOR trial and further 3 publications in CSF and plasma samples^58,60,62,63^.

The cholinergic system (synthesis, esterase activity and receptor density) underlies incompletely understood circadian oscillations, possibly connected to physical activity and stimuli^74,75^. However, we did not examine BCHE FC/activity over time, impeding investigation on circadian regulation.

BCHE’s role goes beyond degrading esters. Elevated activity is linked to obesity, metabolic syndrome, chronic kidney disease and diabetes - all of which are linked to chronic inflammation and oxidative stress^76^. Elevated oxidative stress markers, like malondialdehyde or superoxide dismutase, are observed alongside heightened BCHE activity^76^. It plays complex roles in lipid metabolism by production of precursors of LDL and VLDL, resulting in lipid storage and therefore mitochondrial overload and dysfunction^76^. In contrast, other authors suggest lipolytic properties, involved in lipid membrane loss in neurodegeneration^77^. Also it was linked to hyperhomocysteinaemia, an independent risk factor for neurodegenerative diseases by oxidation, BBB disruption and neuronal cell death^78^. BCHE is involved in regulation of insulin and glucose homeostasis^79^ as well as inactivation of ghrelin by hydrolization^80^. Ghrelin regulates energy intake, modulation of immune response to inflammatory markers as well as memory and learning^81,82^. Therefore, a postoperative decrease of ghrelin may alter cognition. However, BCHE’s metabolic implications should be explored by incorporating metabolomics. BBB disruption, influenced by age, oxidative stress and mitochondrial dysfunction, permits complement and immune cell influx, but remains challenging to measure^12,83^ - no such marker was identified in our cohort.

### Pathway Enrichment Analysis

Our GSEA unravelled complex regulations, most commonly in inflammatory/immune-related pathways, followed by metabolic and hypoxia-related pathways.

#### Inflammatory axis

Both cohorts show postoperative upregulation of many pathways in this domain after surgery (C1-C3). However, pathways in *complement* (2 of 3), *FCERI/FCGR* (8 of 8) and *B cell*s (3 of 3) are specifically enriched in POD patients only. In the cross-sectional approach we can see that 5 *complement* pathways are enriched already at T0, supporting evidence about preoperative (chronic) inflammation in POD. This applies especially for *Activation of C3 and C5* and *Regulation of Complement cascade*. Postoperatively, all but one pathway are significantly enriched (especially Contrast 5), making it difficult to identify immune-related mechanisms *specific* to POD. Yet, overactivation of complement (especially C3 and C5) could be a key driver in pathogenesis - a small mRNA sequencing study revealed significant complement overactivation in POD patients with and without infection^84^. A small MS study in pre- and postoperative immunodepleted CSF showed pathway enrichment of complement (especially C5) and coagulation pathways^85^, further investigation by these authors is underway^86^. The system has been shown to be involved in neurogenesis, neuronal plasticity and neurodegeneration^87^. Additionally, it has been demonstrated that the deposition of complement proteins on neurons triggers phagocytosis, thus causing neurodegeneration and demyelinisation^88,89^. As of now, studies on complement inhibition in POD are lacking. The divergent results of the longitudinal approach highlight the presence of complex immune/inflammatory mechanisms in non-POD patients as well, making it difficult to draw definite conclusions from this approach.

#### Metabolic axis

The longitudinal approach results are heterogeneous. They show strong upregulation of *iron uptake and transport* in POD patients and downregulation in non-PODs. Intracellular iron homeostasis is key for cellular survival - intracellular accumulation leads to ferroptosis, i.e. iron-induced apoptosis via induction of ROS, causing inflammatory cascades and mitochondrial dysfunction^90,91^. It has long been implicated in neuroinflammation and neurodegenerative diseases^90^. Iron overload impairs cellular energy metabolism by hampering the citric acid cycle, increasing anaerobic glycolysis and acidosis^90–92^. The effect of anaesthetics such as sevoflurane, isoflurane and nitrous oxide are complex, numerous and have been described previously: they not only induce ferroptosis but also impede electron transport chain, lead to mitochondrial calcium overload, iron influx imbalance and hamper mitophagia, a process known to degrade dysfunctional mitochondria^93^.

After surgery, POD patients show significantly higher lipid and peptide hormone metabolism than non-POD patients, though only in Contrast 3. Notably, these 2 pathways are among only 4 in the cross-sectional approach that are significantly upregulated in Contrast 6, highlighting their importance. Enhanced lipid and protein metabolism could indicate a compensatory mechanism due to impaired glucose utilization. All other pathways in the cross-sectional approach show downregulation before and after surgery, implying complex metabolic shutdowns prior to surgical trauma. The pathway *Regulation of IGF transport and uptake by IGFBPs* is of special interest as it is downregulated in POD patients (Contrasts 4-6). Insulin-like Growth Factor 1 (IGF-1) plays a crucial role in the pathophysiology of POD and low levels have been consistently associated with POD^12,15,94,95^. IGF-1 supports neuronal regeneration, plasticity, synaptogenesis, and BBB integrity, thus mitigating neuroinflammation and autophagy^95–98^ - all factors relevant to POD. Nevertheless, we did not detect specific downregulation of IGF-1 but IGF-2 (significant in Contrast 1 and 4, insignificant in Contrast 5).

#### Hypoxia axis

GSEA results in the coagulation system, being the only pathways identified as enriched, should be interpreted with caution due to divergence. Fibrin clot formation is reduced in all Contrasts. Non-POD patients show stronger enrichment in *Gamma-carboxylation* pathways after surgery, whereas POD patients display a strong enrichment before surgery compared to non-delirious patients, followed by downregulation - a finding of unknown relevance. There is evidence that POD haemostatic dysregulation is more affecting the thrombocytic axis, as 3 of 4 related pathways show enrichment in Contrast 6. Also, there is significant upregulation of *Cell surface interactions at the vascular wall*, supporting thrombocytic interaction with endothelium that might result in microthrombosis as well as proinflammatory signalling^99,100^. Moreover, thrombocyte activation is a frequent event in sepsis and may play a role in the increased incidence of delirium observed in septic patients^101,102^. In the review by Wiredu et al., 3 out of the 10 top GO terms from 370 suspected POD markers were *platelet*-associated, all of which show activation in our study. This supports our data quality but also underlines thrombocytes’ involvement in POD^16^. The review covers 8 studies investigating POD-associated proteins in 484 patients and functional analysis of 370 proteins revealed the top 10 GO terms: 3 linked to the immune system, 5 to haemostasis/platelet function and 2 to metabolic pathways^16^. GO terms of the review in relation to our GSEA can be found in Tab. S10. In 8 of the 10 terms we found enrichment in Contrasts 4-6, in one term we found enrichment in Contrasts 1-3 and in one we found upregulation in all Contrasts (*Immune System*). This highlights the concordance of our profiling with previous works and supports current hypotheses that most alterations likely will be found in the domains of *inflammation* and *haemostasis*. Han et al. performed Reactome enrichment of 15 preoperative CSF samples - our analysis is enriched in 7 of their 15 top terms. This suggests certain overlaps between CSF and plasma proteomic signatures, giving reason for parallel CSF and plasma investigation.

### Exploratory Proteomic Classification Analysis

To our knowledge, no proteomic classification approach investigating POD-related profiles has been reported to date^12,16^. However, Tripp et al. described an 11-metabolite signature with an AUC of 0.838 using targeted MS in plasma samples from 52 POD patients in a matched case-control design^103^. Integration of metabolomic and proteomic MS efforts may enhance future biological characterisation of POD-associated patterns. Notably, BCHE levels emerged as a key feature, highlighting the complex involvement of cholinergic pathways^76,77^. Remarkably, 4 of the 8 proteins were variable light or heavy chains of immunoglobulins, whose plasma levels are highly variable and influenced by numerous factors (e.g. age, sex, lifestyle, BMI)^104^. Also here, immunodepletion could have altered their detection. Due to limited information available on IGKV3-11, IGHV3-15 and IGLV3-27, their implications remain unclear. Factor V (F5) was the second most differentiating protein, with mild but significant downregulation in POD patients. Similarly, Han et al. have shown low F5 levels in preoperative CSF to correlate with POD emergence, suggesting a possible role in amyloid β destabilisation^105^.

Further classification analyses using linear discriminant analysis, support vector machines, glmnet approaches and neural network modeling did not yield superior discriminative performance compared to LR. The presented classification, derived from a clinically diverse cohort within a matched case-control design, limits extrapolation to general population-based settings. Replication in larger sample batches is required to validate observed mechanistic patterns.

### Strengths and Limitations

Our study provides insight into various mechanisms by employing two approaches and GSEA in a contrasting design. Also, we present one of the largest case-control cohorts for MS-based POD proteomics to date. Our data quality is supported by using paired, balanced sample sets. However, we acknowledge limitations to our study, including its single-center and retrospective nature as well as its matched case-control design. Consequently, the findings cannot be generalised beyond the characteristics of our specific cohort. Furthermore, the cohort was heterogeneous in terms of history, POD subtype and severity, surgical procedure, DoA and preoperative performance. We deliberately avoided restricting the study pipeline to highly specific patient subgroups in order to reflect generalised perioperative changes across a broader patient population. Moreover, plasma sampling times were not standardised, potentially affecting proteins underlying circadian rhythms^74^. Finally, no orthogonal, independent affinity-based validation (e.g., ELISA) or absolute quantification of proteins was performed, as antibody availability for most candidates is limited. Previous publications partially addressed validation but only for selected targets^16,44,106–108^.

## Conclusions

HT-LCMS and functional proteomics are valuable tools for investigating pathophysiological mechanisms underlying POD, with the potential to i) enhance our comprehension of its complex pathogenesis, ii) generate novel mechanistic hypotheses and iii) identify proteomic patterns associated with perioperative vulnerability. Incorporating additional OMICS approaches, particularly metabolomics, may enrich mechanistic insights. Our findings require reproduction in larger, independent cohorts and secondary validation by orthogonal technologies to strengthen the biological relevance of the observed patterns.

## Declarations

### Data Availability Statement

The dataset analysed for and during the presented study are available from the corresponding author upon reasonable request. The procedure of sample selection and creation of a balanced and matched sample set is available in the Supplementary Material.

## Acknowledgements

We thank our team of investigators, medical doctoral students and study nurses: Rudolf Mörgeli, Friedrich Borchers, Anika Müller, Alissa Wolf, Fatima Yürek, Daniel Hadzidiakos, Florian Lammers-Lietz, Ilse Kant, Simone van Montfort, Gunnar Lachmann, Anika Alon, Sina Rosenblender, Tuba Aslan, Markus Laubach, Felix Müller, Emmanuel Keller, Eleftheria Papadaki, Saya Speidel, Bennet Borak, Steffi Herferth, Johannes Lange, Helene Michler, Juliane Dörfler, Anton Jacobshagen, Petra Kozma, Marinus Fislage, Wolf Rüdiger Brockhaus, Luisa Rothe, Pola Neuling, Ken-Dieter Michel, Firas Nosirat, Maryam Kurpanik, Sophia Kuenz, Lukas Roediger, Irene Mergele, Leopold Rupp, Marie Graunke, and Victoria Windmann. We thank Jochen Kruppa and Janine Wiebach for the design of the plate loading and Birgit Brandt for the processing of samples. We thank all staff members of the HT-MS Charité Core Facility and the Department of Biochemistry for sample and data processing.

## Author Contributions (according to CRediT)

**Mario Lamping**: Investigation, Resources, Data Curation, Writing - Original Draft, Writing - Review & Editing

**Maria Heinrich**: Conceptualisation, Methodology, Formal Analysis, Investigation, Resources, Data Curation, Writing - Original Draft

**Vadim Farztdinov**: Conceptualisation, Methodology, Software, Validation, Formal analysis, Data Curation, Writing - Review & Editing, Visualisation

**Clarissa von Haefen**: Resources, Writing - Original Draft, Writing - Review & Editing

**Jayanth Sreekanth**: Writing - Review & Editing

**Michael Mülleder**: Conceptualisation, Methodology, Software, Validation, Formal Analysis, Investigation, Resources, Data Curation, Writing - Review & Editing, Supervision

**Markus Ralser**: Conceptualisation, Resources, Supervision, Project administration, Funding acquisition

**Georg Winterer**: Conceptualisation, Investigation, Resources, Writing - Review & Editing, Funding acquisition

**Claudia D. Spies**: Conceptualisation, Methodology, Investigation, Resources, Writing - Review & Editing, Supervision, Project administration, Funding acquisition

## List of Abbreviations

ABC: Ammonium Bicarbonate
ACh: Acetylcholine
ADL: Activities of Daily Living
APCS: Serum Amyloid P Component
APOA4: Apolipoprotein A-IV
APOF: Apolipoprotein F
ASA PS: American Society of Anaesthesiologists Physical Status
AUC: Area Under the Curve
B2M: Beta-2-Microglobulin
BBB: Blood-Brain Barrier
BCHE: Butyrylcholinesterase
BMI: Body Mass Index
C18orf63: Chromosome 18 Open Reading Frame 63
C1R: Complement Component 1, R Subcomponent
C2: Complement Component 2
C3: Complement Component 3
C4: Complement Component 4
C5: Complement Component 5
CAM-ICU: Confusion Assessment Method for the Intensive Care Unit
CCI: Charlson Comorbidity Index
CFHR2: Complement Factor H-Related Protein 2
CFHR4: Complement Factor H-Related Protein 4
ChE: Cholinesterase
CNDP1: Carnosine Dipeptidase 1
CRP: C-Reactive Protein
CSF: Cerebrospinal Fluid
CST3: Cystatin C
DE0: non-POD patients
DE1: POD patients
DIA-NN: Data-Independent Acquisition Neural Networks
SWATH: Sequential Window Acquisition of All Theoretical Mass Spectra
DoA: Duration of Anaesthesia
DSM-5: Diagnostic and Statistical Manual of Mental Disorders, 5th Edition
EEG: Electroencephalogram
ELISA: Enzyme-Linked Immunosorbent Assay
F5: Factor V
FBLN1: Fibulin 1
FC: Fold Change
FCGBP: Fc Fragment of IgG Binding Protein
FCGR: Fc Gamma Receptor
FCERI: Fc Epsilon Receptor I
FDR: False Discovery Rate
FGA: Fibrinogen Alpha Chain
GO: Gene Ontology
GOBP: Gene Ontology Biological Process
GPX3: Glutathione Peroxidase 3
GSEA: Gene Set Enrichment Analysis
HB: Haemoglobin
HBA1: Haemoglobin Subunit Alpha 1
HBD: Haemoglobin Subunit Delta
HT-LCMS: High-Throughput Liquid Chromatography-Mass Spectrometry
IGF-1: Insulin-Like Growth Factor 1
IGF2: Insulin-Like Growth Factor 2
IGFALS: Insulin-Like Growth Factor Binding Protein, Acid-Labile Subunit
IGFBPs: Insulin-Like Growth Factor Binding Proteins
IGHV: Immunoglobulin Heavy Variable
IGKV: Immunoglobulin Kappa Variable
IL: Interleukin
iNOS: inducible nitric oxide synthase
LBP: Lipopolysaccharide-Binding Protein
LPA: Lipoprotein(a)
LR: Logistic Regression
LYZ: Lysozyme
MAD: Median Absolute Deviation
MASP1: Mannose-Associated Serine Protease 1
MBL2: Mannose-Binding Lectin 2
MMSE: Mini-Mental State Examination
MNA: Mini Nutritional Assessment
MST1: Macrophage Stimulating 1
NCD: Neurocognitive Disorder
NES: Normalised Enrichment Score
Nu-DESC: Nursing Delirium Screening Scale
PCA: Principal Component Analysis
PGLYRP2: Peptidoglycan Recognition Protein 2
PLM: Panel Linear Model
POCD: Postoperative Cognitive Dysfunction
POD: Postoperative Delirium
ppm: Parts Per Million
PROZ: Protein Z
QSOX1: Quiescin Sulfhydryl Oxidase 1
ROS: Reactive Oxygen Species
S100A9: S100 Calcium-Binding Protein A9
S100B: S100 Calcium-Binding Protein B
SAA1: Serum Amyloid A1
SD: Standard Deviation
SELL: L-Selectin
SERPIND1: Serpin Family D Member 1
SWATH: Sequential Window Acquisition of All Theoretical Mass Spectra
T0: Preoperative Time Point 0 (Baseline)
T1: Postoperative Time Point 1 (Day 1)
TFRC: Transferrin Receptor
TMSB4X: Thymosin Beta-4 X-Linked
TNF-ɑ: Tumor Necrosis Factor-Alpha
VTN: Vitronectin

