## Supplementary Material for "Mechanistic Insights Into Postoperative Delirium Using Untargeted High-Throughput Proteomics in Elderly Patients - A Case-Control Study"

### Figures

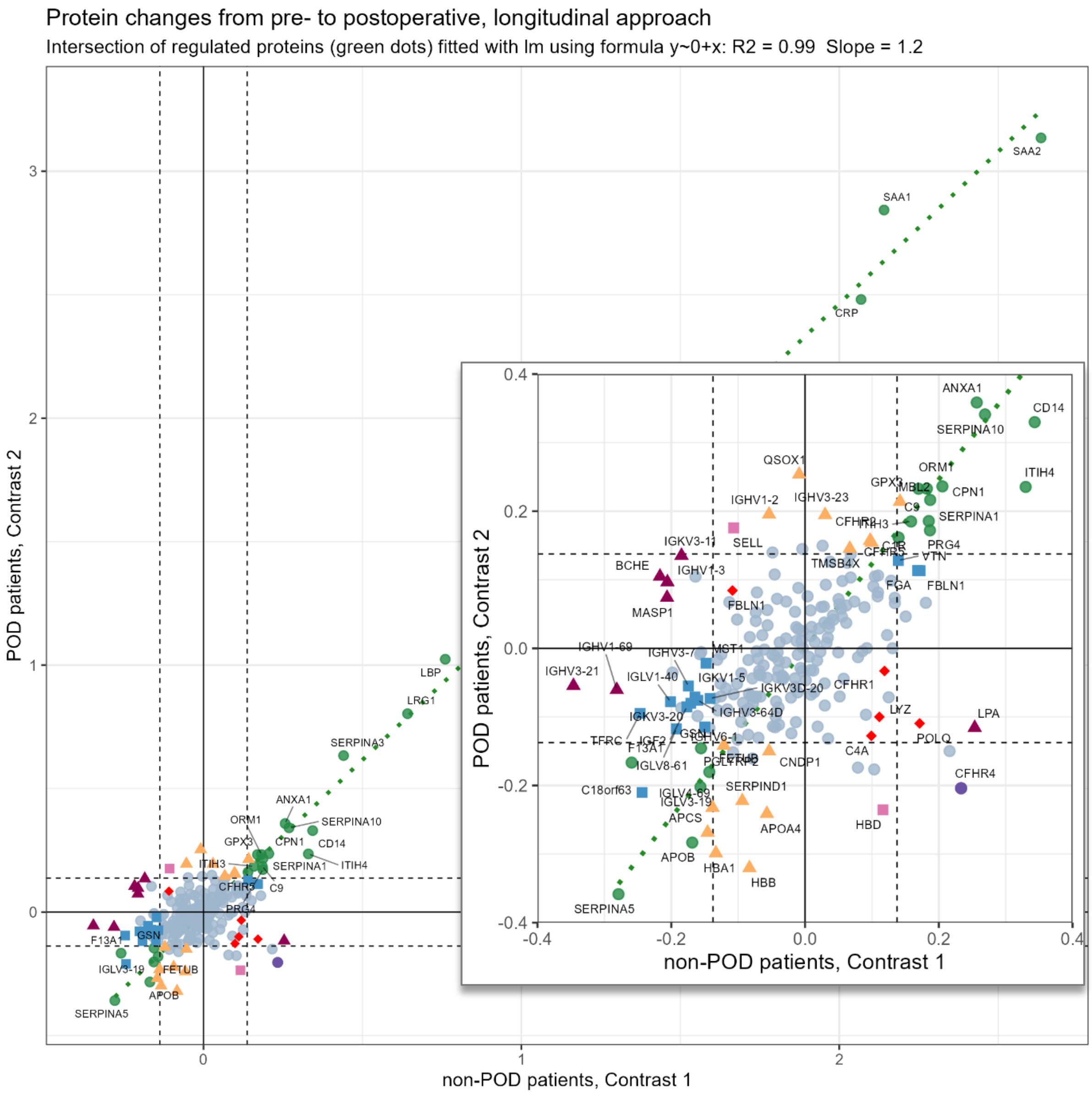

**Fig. S1** - Scatterplot of log2(FC) in the longitudinal approach in Contrast 1 (non-POD patients, x-axis) and Contrast 2 (POD patients, y-axis). Green dots mark the proteins significantly regulated in both contrasts. The insert shows a magnified part of the figure around the origin.

Blue = only regulated in Contrast 1: 15 proteins
Orange = only regulated in Contrast 2: 15 proteins
Green = regulated in Contrasts 1 and 2: 24 proteins
Red = only regulated in Contrast 3: 5 proteins
Violet = regulated in Contrasts 2 and 3: 2 proteins
Dark Violet = regulated in Contrasts 1 and 3: 7 proteins
Pink = regulated in Contrasts 1, 2 and 3: 1 protein
Grey = not regulated

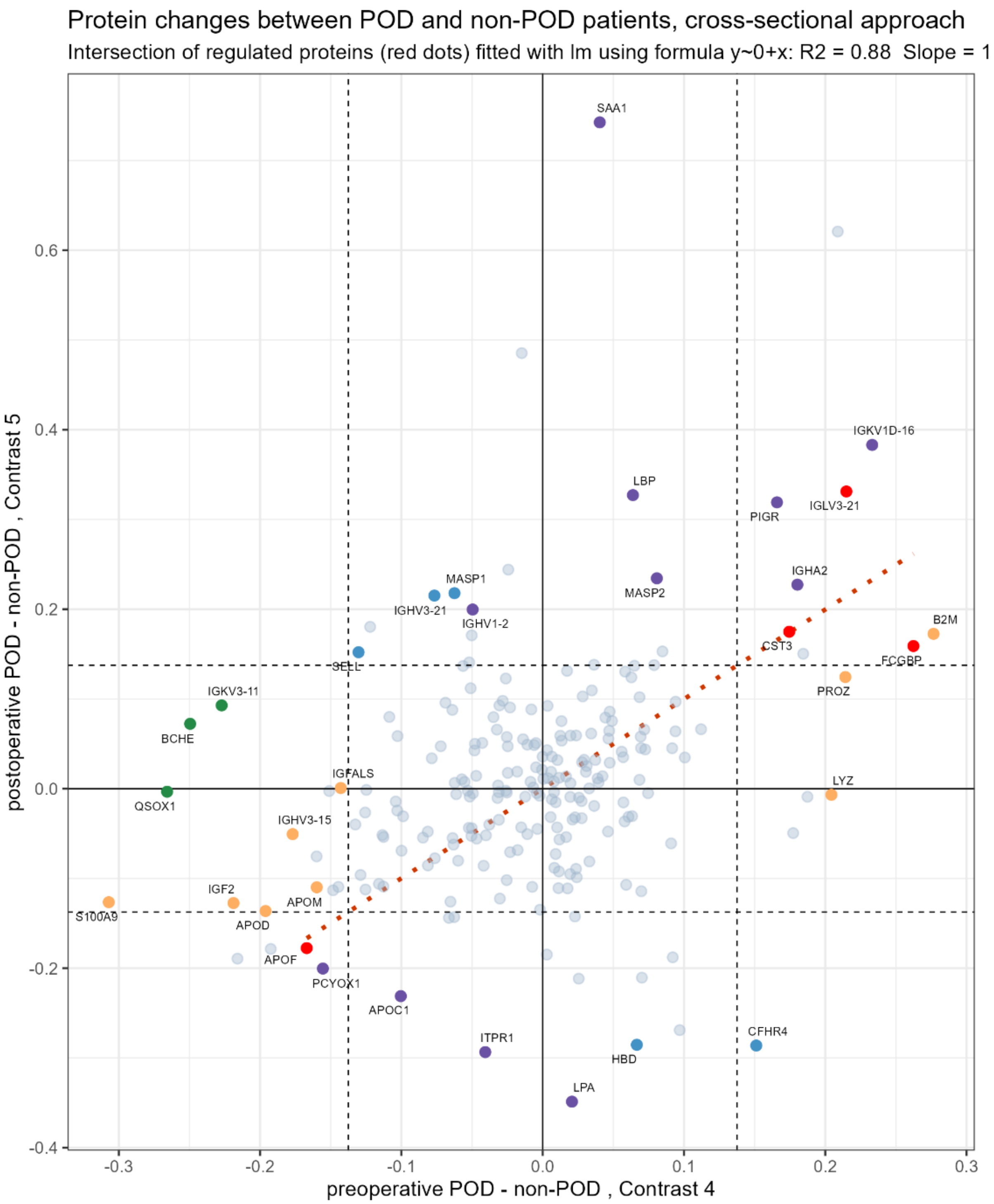

**Fig. S2** - Scatterplot of log2(FC) in the cross-sectional approach in Contrast 4 (preoperative, x-axis) and Contrast 5 (postoperative, y-axis). Red dots mark the proteins significantly regulated in both contrasts.
Orange = only regulated in Contrast 4: 9 proteins
Pink = only regulated in Contrast 5: 11 proteins
Red = regulated in Contrasts 4 and 5: 4 proteins
Blue = only regulated in Contrasts 5 and 6: 5 proteins
Green = only regulated in Contrasts 4 and 6: 3 proteins
Grey = not regulated

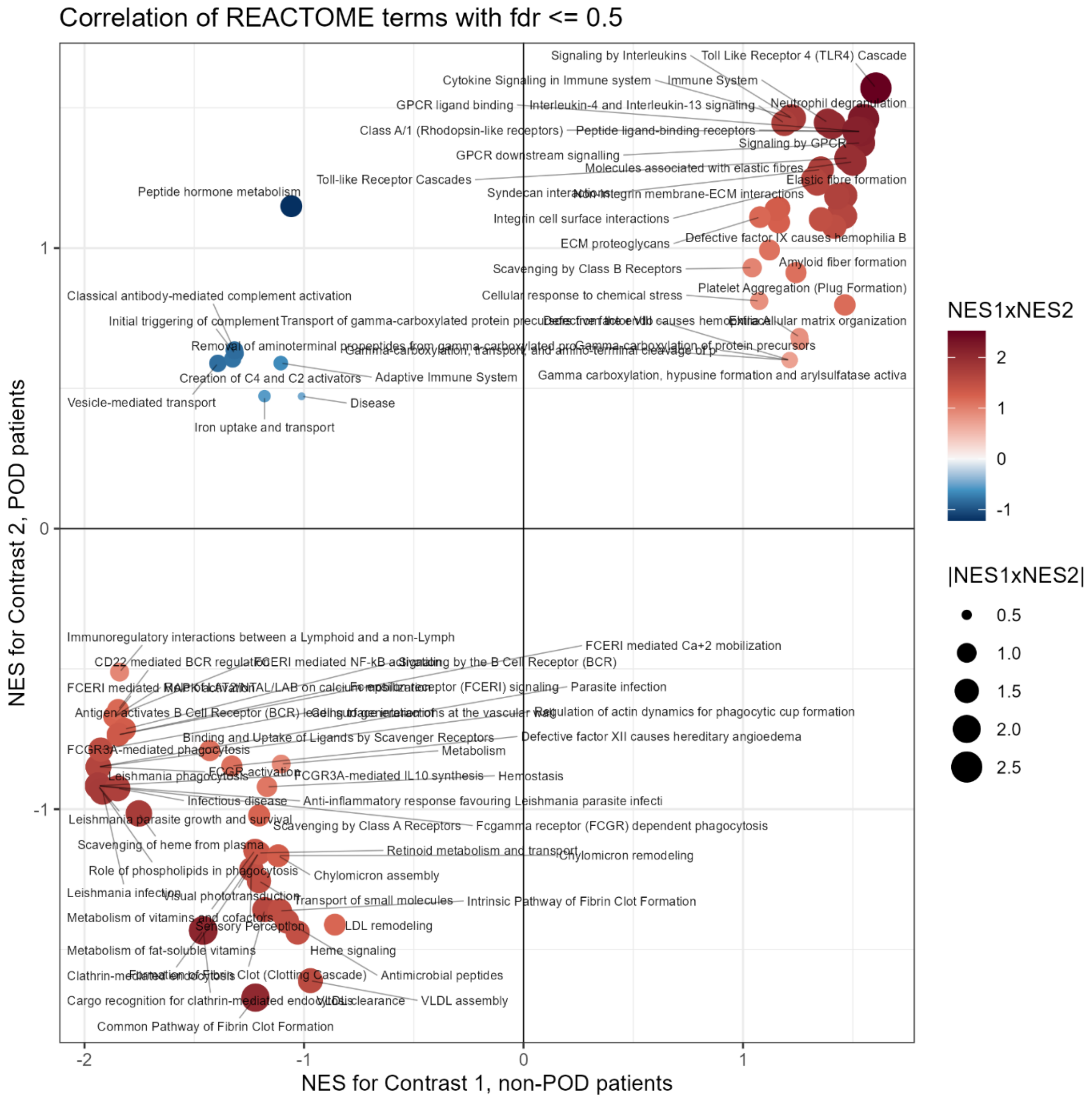

**Fig. S3** – Scatterplot of REACTOME Pathway Enrichment for the longitudinal approach, showing enrichment (NES) in Contrast 1 (x-axis) and Contrast 2 (y-axis) with fdr ≤0.5. NES = normalised enrichment score.

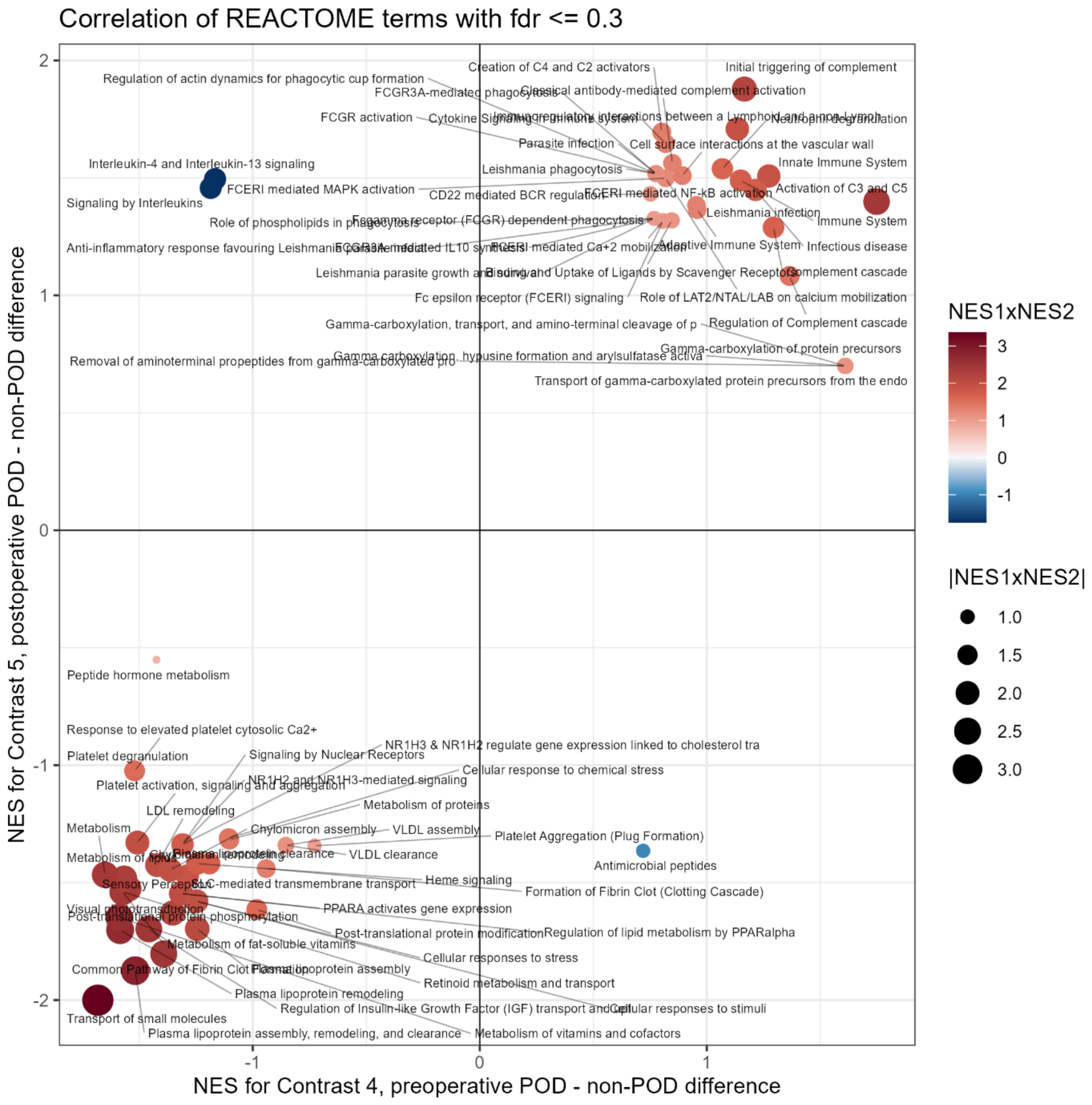

**Fig. S4** – Scatterplot of REACTOME Pathway Enrichment for the cross-sectional approach, showing enrichment (NES) in Contrast 4 (x-axis) and Contrast 5 (y-axis) with fdr ≤0.3. NES = normalised enrichment score.

### Tables

**Tab. S1** – Total Dataset

| Subgroup Diagnosis / Visit | N Samples | N Duplicates | Total |
|---|---|---|---|
| DE0_T0 | 112 | 5 | 117 |
| DE0_T1 | 96 | 7 | 103 |
| | 70 | | 70 |
| DE1_T0 | 112 | 10 | 122 |
| DE1_T1 | 100 | 6 | 106 |
| DE1_T2 | 41 | | 41 |
| (NA) | 1 | | 1 |
| **Total** | **532** | **28** | **560** |

Number of samples in each disease visit subgroup before removing quality outliers. One sample (BIM349) was without clinical data. DE0 = no delirium, DE1 = delirium, T0 = preoperative, T1 = postoperative day 1, T2 = postoperative day 2, NA = not available

**Tab. S2** – Binned Samples

| Diagnosis/Visit | T0 | T1 | Total |
|---|---|---|---|
| **DE0** | **110** | **93** | **203** |
| Not @ both visits | 21 | 4 | 25 |
| @ both visits | 89 | 89 | 178 |
| **DE1** | **112** | **100** | **212** |
| Not @ both visits | 16 | 4 | 20 |
| @ both visits | 96 | 96 | 192 |
| **Total** | **222** | **193** | **415** |

Number of samples in each group after removing quality outliers, T2 and duplicates. Not all patients had sampling at both visits. DE0 = no delirium, DE1 = delirium, T0 = preoperative, T1 = postoperative day 1.

**Tab. S3** – Samples According to MNA Status

| Nutrition Status | MNA1 | MNA2 | MNA3 | MNA9 | Total |
|---|---|---|---|---|---|
| **DE0** | **7** | **20** | **55** | **2** | **84** |
| female | 3 | 11 | 27 | 1 | 42 |
| male | 4 | 9 | 28 | 1 | 42 |
| **DE1** | **8** | **24** | **51** | **1** | **84** |
| female | 5 | 12 | 24 | 1 | 42 |
| male | 3 | 12 | 27 | | 42 |
| **Total** | **15** | **44** | **106** | **3** | **168** |

Number of samples in each disease subgroup according to nutritional status. Values at T0 and T1 are the same due to balancing. DE0 = no delirium, DE1 = delirium, MNA = Mini Nutritional Assessment. MNA9 stands for missing information on nutritional status.

**Tab. S4** – Age According to MNA

| Nutrition Status | MNA1 | MNA2 | MNA3 | MNA9 | Total Avg |
|---|---|---|---|---|---|
| **DE0** | **72** | **73** | **73** | **73** | **73** |
| female | 75 | 72 | 74 | 76 | 74 |
| male | 71 | 74 | 72 | 70 | 72 |
| **DE1** | **75** | **74** | **74** | **75** | **74** |
| female | 75 | 72 | 74 | 75 | 74 |
| male | 74 | 76 | 73 | | 74 |
| **Total Avg** | **74** | **74** | **73** | **74** | **73** |

Average (Avg) age of patient in each disease subgroup (years). Values at T0 and T1 are the same due to balancing. DE0 = no delirium, DE1 = delirium, MNA = Mini Nutritional Assessment. MNA9 stands for missing information on nutritional status.

**Tab. S1** - Initial Distribution of Samples Over ASA and Duration of Anaesthesia

| | DE0 | DE1 | all |
|---|---|---|---|
| **ASA_1** | | | |
| No of samples | 2 | 3 | 5 |
| Average Age | 68 | 72 | 70 |
| Average DoA | 75 | 259 | 185 |
| **ASA_2** | | | |
| No of samples | 73 | 51 | 124 |
| Average Age | 73 | 73 | 73 |
| Average DoA | 238 | 301 | 264 |
| **ASA_3** | | | |
| No of samples | 36 | 58 | 94 |
| Average Age | 73 | 75 | 74 |
| Average DoA | 247 | 350 | 310 |
| **ASA_4** | | | |
| No of samples | 1 | | 1 |
| Average Age | 71 | | 71 |
| Average DoA | 86 | | 86 |
| **Total:** all samples | **112** | **112** | **224** |
| **Total:** Average Age | **73** | **74** | **73** |
| **Total:** Average DoA | **236** | **325** | **281** |

**Tab. S2** – Balanced Set of Samples Over ASA and Duration of Anaesthesia

| | DE0 | DE1 | all |
|---|---|---|---|
| **ASA_1** | | | |
| No of samples | 1 | 2 | 3 |
| Average Age | 69 | 72 | 71 |
| Average DoA | 52 | 332 | 238 |
| **ASA_2** | | | |
| No of samples | 57 | 37 | 94 |
| Average Age | 73 | 73 | 73 |
| Average DoA | 247 | 314 | 274 |
| **ASA_3** | | | |
| No of samples | 26 | 45 | 71 |
| Average Age | 73 | 75 | 74 |
| Average DoA | 273 | 357 | 326 |
| **Total:** all samples | **84** | **84** | **168** |
| **Total:** Average Age | **73** | **74** | **73** |
| **Total:** Average DoA | **253** | **337** | **295** |

Tab. S5 shows confounding between POD, ASA and DoA at T0. Tab. S6 shows altered distribution over ASA and DoA after balancing. In this case, the distribution of samples is the same at T0 and T1. There was only marginal improvement in the number of samples over ASA status and DoA between POD and non-POD patients. ASA PS = physical status according to the American Society of Anaesthesiologists, DE0 = no delirium, DE1 = delirium, DoA = duration of anaesthesia, T0 = preoperative, T1 = postoperative day 1.

**Tab. S7** – Reactome Pathway Enrichment (Longitudinal Approach)

| Description Pathway | Set Size | NES | p-value | q-value | Core enrichment | Category |
|---|---|---|---|---|---|---|
| **Immunoregulatory interactions between a Lymphoid and a non-Lymphoid cell** | 25 | 1.62 | 0.01 | 0.25 | SELL/IGKV3-11/IGHV1-69/IGHV1-2/IGKV4-1/IGHV2-5/IGHV3-7/IGHV3-23/IGHV3-13/IGKV1-5/IGLV3-21/IGKV3-20/IGLV2-8/IGKV1D-16 | REAC |
| **CD22 mediated BCR regulation** | 23 | 1.58 | 0.02 | 0.25 | IGKV3-11/IGHV1-69/IGHV1-2/IGKV4-1/IGHV2-5/IGHV3-7/IGHV3-23/IGHV3-13/IGKV1-5/IGLV3-21/IGKV3-20/IGLV2-8/IGKV1D-16 | REAC |
| **LDL remodeling** | 3 | -1.55 | 0.02 | 0.25 | APOB/LPA | REAC |
| **Role of LAT2/NTAL/LAB on calcium mobilization** | 22 | 1.57 | 0.02 | 0.25 | IGKV3-11/IGHV1-69/IGHV1-2/IGKV4-1/IGHV2-5/IGHV3-7/IGHV3-23/IGHV3-13/IGKV1-5/IGLV3-21/IGKV3-20/IGLV2-8/IGKV1D-16 | REAC |
| **Creation of C4 and C2 activators** | 32 | 1.55 | 0.02 | 0.25 | IGKV3-11/MASP1/IGHV1-69/IGHV1-2/IGKV4-1/IGHV2-5/IGHV3-7/IGHV3-23/IGHV3-13/IGKV1-5/IGLV3-21/IGKV3-20/IGLV2-8/IGKV1D-16/IGLV1-40/CRP | REAC |
| **FCGR3A-mediated phagocytosis** | 25 | 1.55 | 0.02 | 0.25 | IGKV3-11/IGHV1-69/IGHV1-2/IGKV4-1/IGHV2-5/IGHV3-7/IGHV3-23/IGHV3-13/IGKV1-5/IGLV3-21/IGKV3-20/IGLV2-8/IGKV1D-16 | REAC |
| **Vesicle-mediated transport** | 42 | 1.54 | 0.03 | 0.28 | IGKV3-11/MASP1/IGHV1-69/IGHV1-2/IGKV4-1/IGHV2-5/IGHV3-7/IGHV3-23/TF/SAA1/TFRC/APOE/IGHV3-13/IGKV1-5/IGLV3-21/JCHAIN/IGKV3-20/ALB/IGLV2-8/IGKV1D-16 | REAC |
| **Adaptive Immune System** | 33 | 1.49 | 0.03 | 0.28 | SELL/IGKV3-11/IGHV1-69/IGHV1-2/IGKV4-1/IGHV2-5/IGHV3-7/IGHV3-23/IGHV3-13/IGKV1-5/IGLV3-21/IGKV3-20/S100A9/IGLV2-8/IGKV1D-16 | REAC |
| **Peptide hormone metabolism** | 3 | 1.45 | 0.03 | 0.28 | BCHE | REAC |
| **Intrinsic Pathway of Fibrin Clot Formation** | 15 | -1.55 | 0.03 | 0.28 | SERPINA5/A2M/PROC/PROS1/F11/F2/SERPIND1 | REAC |
| **Binding and Uptake of Ligands by Scavenger Receptors** | 38 | 1.45 | 0.04 | 0.28 | IGKV3-11/MASP1/IGHV1-69/IGHV1-2/IGKV4- | REAC |

| | | | | | | |
|---|---|---|---|---|---|---|
| | | | | | 1/IGHV2-5/IGHV3-7/IGHV3-23/SAA1/APOE/IGHV3-13/IGKV1-5/IGLV3-21/JCHAIN/IGKV3-20/ALB/IGLV2-8/IGKV1D-16 | |
| **Classical antibody-mediated complement activation** | 29 | 1.49 | 0.04 | 0.28 | IGKV3-11/IGHV1-69/IGHV1-2/IGKV4-1/IGHV2-5/IGHV3-7/IGHV3-23/IGHV3-13/IGKV1-5/IGLV3-21/IGKV3-20/IGLV2-8/IGKV1D-16/IGLV1-40/CRP | REAC |
| **Common Pathway of Fibrin Clot Formation** | 13 | -1.58 | 0.04 | 0.28 | FGB/FGG/SERPINA5/PROC/PROS1/F2/SERPIND1 | REAC |
| **Initial triggering of complement** | 38 | 1.42 | 0.05 | 0.28 | IGKV3-11/MASP1/IGHV1-69/IGHV1-2/CFB/IGKV4-1/IGHV2-5/IGHV3-7/IGHV3-23/IGHV3-13/IGKV1-5/IGLV3-21/IGKV3-20/IGLV2-8/IGKV1D-16/C2/IGLV1-40/CRP | REAC |
| **Metabolism of lipids** | 9 | 1.49 | 0.05 | 0.28 | BCHE | REAC |
| **Signaling by the B Cell Receptor (BCR)** | 24 | 1.47 | 0.05 | 0.28 | IGKV3-11/IGHV1-69/IGHV1-2/IGKV4-1/IGHV2-5/IGHV3-7/IGHV3-23/IGHV3-13/IGKV1-5/IGLV3-21/IGKV3-20/IGLV2-8/IGKV1D-16 | REAC |
| **FCERI mediated Ca+2 mobilization** | 23 | 1.47 | 0.05 | 0.28 | IGKV3-11/IGHV1-69/IGHV1-2/IGKV4-1/IGHV2-5/IGHV3-7/IGHV3-23/IGHV3-13/IGKV1-5/IGLV3-21/IGKV3-20/IGLV2-8/IGKV1D-16 | REAC |
| **Role of phospholipids in phagocytosis** | 26 | 1.43 | 0.06 | 0.28 | IGKV3-11/IGHV1-69/IGHV1-2/IGKV4-1/IGHV2-5/IGHV3-7/IGHV3-23/IGHV3-13/IGKV1-5/IGLV3-21/IGKV3-20/IGLV2-8/IGKV1D-16 | REAC |
| **Heme signaling** | 4 | -1.51 | 0.06 | 0.28 | HBA1/HBB/APOB | REAC |
| **Cell surface interactions at the vascular wall** | 34 | 1.39 | 0.07 | 0.28 | SELL/IGKV3-11/IGHV1-69/IGHV1-2/IGKV4-1/IGHV2-5/IGHV3-7/IGHV3-23/IGHV3-13/IGKV1-5/IGLV3-21/JCHAIN/IGKV3-20/IGLV2-8/IGKV1D-16 | REAC |
| **Gamma-carboxylation, transport, and amino-terminal cleavage of proteins** | 6 | -1.48 | 0.08 | 0.28 | F9/PROZ/PROC/PROS1/F2 | REAC |
| **Iron uptake and transport** | 3 | 1.38 | 0.08 | 0.28 | TF/TFRC | REAC |

Reactome pathways enriched with FDR ≤0.3 in the longitudinal approach with pure POD effect (Contrast 3). NES – normalised enrichment score.

**Tab. S8** - Reactome and GOBP Pathway Enrichment (Cross-sectional Approach)

| Description Pathway | Set Size | NES | p-value | q-value | Core enrichment | Category |
|---|---|---|---|---|---|---|
| **Immunoregulatory interactions between a Lymphoid and a non-Lymphoid cell** | 25 | 1.5 | 0.032 | 0.41 | SELL/IGKV3-11/IGHV1-2/IGHV1-69/IGHV2-5/IGKV4-1/IGHV3-23/IGHV3-7/IGHV3-13/IGLV3-21/IGKV1D-16/IGKV1-5/IGKV3-20 | REAC |
| **Creation of C4 and C2 activators** | 32 | 1.5 | 0.036 | 0.41 | MASP1/IGKV3-11/IGHV1-2/IGHV1-69/IGHV2-5/IGKV4-1/IGHV3-23/IGHV3-7/IGHV3-13/CRP/IGLV3-21/IGKV1D-16/IGKV1-5/IGKV3-20 | REAC |
| **Peptide hormone metabolism** | 3 | 1.4 | 0.046 | 0.41 | BCHE | REAC |
| **Initial triggering of complement** | 38 | 1.4 | 0.048 | 0.41 | MASP1/IGKV3-11/IGHV1-2/IGHV1-69/IGHV2-5/CFB/IGKV4-1/IGHV3-23/IGHV3-7/IGHV3-13/CRP/IGLV3-21/IGKV1D-16/C2/IGKV1-5 | REAC |
| **Vesicle-mediated transport** | 42 | 1.4 | 0.057 | 0.41 | MASP1/IGKV3-11/IGHV1-2/IGHV1-69/IGHV2-5/SAA1/IGKV4-1/IGHV3-23/TFRC/TF/IGHV3-7/IGHV3-13/IGLV3-21/ALB/IGKV1D-16/IGKV1-5/APOE/IGKV3-20 | REAC |
| **Neutrophil degranulation** | 24 | 1.2 | 0.273 | 0.66 | SELL/A1BG/QSOX1/SERPINA3/ORM2/S100A9/LRG1 | REAC |
| **Sensory Perception** | 10 | -1.2 | 0.286 | 0.66 | APOC3/TTR/APOB/APOA4 | REAC |
| **Platelet degranulation** | 38 | 1.1 | 0.390 | 0.71 | A1BG/QSOX1/SERPINA3/ORM2/CLEC3B/F13A1/TF/VWF/SERPINA4/ALB/TMSB4X/SERPING1/IGF2/ITIH4 | REAC |
| **hydrogen peroxide metabolic process** | 6 | -1.7 | 0.009 | 0.94 | HBA1/HP/HBB/APOA4/HBD | GOBP |
|  | 37 | 1.5 | 0.025 | 0.94 | IGHV3-21/IGHV1-3/IGHV1-2/IGHV1-69/IGHV2-5/LBP/IGHV3-23/IGHV3-15/TFRC/ANXA1/IGHV3-7/IGHV3-13 | GOBP |
| **cell communication** | 81 | 1.4 |  | 0.94 | BCHE/IGHV3-21/IGHV1-3/IGHV1-2/FBLN1/IGHV1-69/IGHV2-5/IGFALS/LBP/IGHV3-23/MST1/IGHV3-15/TFRC/TF/S100A9/LRG1/VWF/ANXA1/IGHV3-7/IGHV3-13/ALB/IGHV2-26/APOE/TMSB4X/IGF2/PIGR/IGHV3-72/APOD | GOBP |
| **behavior** | 7 | 1.5 | 0.044 | 0.94 | BCHE | GOBP |

| | | | | | | |
|---|---|---|---|---|---|---|
| **extracellular matrix organization** | 9 | 1.6 | 0.046 | 0.94 | QSOX1/FBLN1/LUM/TGFBI | GOBP |
| **regulation of trans-synaptic signaling** | 3 | 1.4 | 0.047 | 0.94 | BCHE | GOBP |
| **lipid transport** | 24 | -1.5 | 0.047 | 0.94 | PON1/SERPINA5/APOL1/APOC1/APOB/CLU/APOA4/LPA/CFHR4 | GOBP |
| **cell recognition** | 36 | 1.4 | 0.047 | 0.94 | IGHV3-21/IGHV1-3/IGHV1-2/IGHV1-69/IGHV2-5/LBP/IGHV3-23/IGHV3-15/IGHV3-7/IGHV3-13/CRP | GOBP |

Reactome (REAC) and GOBP terms enriched with p-value ≤0.05 in the cross-sectional approach with pure POD effect (Contrast 6). Enrichment has high FDR >0.4. NES – normalised enrichment score.

**Tab. S9** – Generalised Linear Model Results For 8 Proteins Associated With POD Classification

| **Protein** | **Estimate** | **Std. Error** | **z-value** | **Pr(>\|z\|)** | **Significance** |
|---|---|---|---|---|---|
| BCHE | -28.396 | 0.6239 | -4.551 | 5.33e-06 | *** |
| F5 | -48.650 | 11.073 | -4.393 | 1.12e-05 | *** |
| IGKV3-11 | -18.714 | 0.4880 | -3.835 | 0.000126 | *** |
| IGLV3-27 | 11.545 | 0.3386 | 3.410 | 0.000651 | *** |
| PROZ | 15.553 | 0.4540 | 3.425 | 0.000614 | *** |
| IGHV3-15 | -20.796 | 0.5478 | -3.796 | 0.000147 | *** |
| QSOX1 | -13.345 | 0.3812 | -3.501 | 0.000464 | *** |
| IGLV3-21 | 10.874 | 0.3629 | 2.997 | 0.002730 | ** |

Significance codes: 0 '***' 0.001 '**' 0.01 '*'

**Tab. S10** – Enrichment of 10 Pathways Compared to Wiredu et al.

| **Pathway Description** | **C1** | **C2** | **C3** | **C4** | **C5** | **C6** |
|---|---|---|---|---|---|---|
| **Immune System** | + | + | + | + | + | + |
| **Hemostasis** | - | - | + | n.e. | n.e. | n.e. |
| **Cytokine signaling in Immune System** | n.e. | n.e. | n.e. | (+) | + | + |
| **Signaling by Interleukins** | n.e. | n.e. | n.e. | - | + | + |
| **Platelet activation, signaling and aggregation** | n.e. | n.e. | n.e. | - | - | + |
| **Response to elevated platelet cytosolic Ca2+** | n.e. | n.e. | n.e. | - | - | + |
| **Platelet degranulation** | n.e. | n.e. | n.e. | - | - | + |
| **Regulation of IGF transport and uptake by IGFBPs** | n.e. | n.e. | n.e. | - | - | - |
| **Post translational protein phosphorylation** | n.e. | n.e. | n.e. | - | - | - |
| **Formation of Fibrin Clot** | n.e. | n.e. | n.e. | - | - | - |

Wiredu et al. published the top 10 GO terms in a systematic review of 8 studies exploring alterations and markers in POD patients (red = inflammatory/immune axis, blue = haemostasis axis, green = metabolic axis). This table displays the enrichments observed in our GSEA on the longitudinal approach (C1-3) and the cross-sectional approach (C4-6). + = upregulated, - = downregulated, (+) = not significant, n.e. = not enriched.
